# Blueprint for a Pop-up SARS-CoV-2 Testing Lab

**DOI:** 10.1101/2020.04.11.20061424

**Authors:** Innovative Genomics Institute SARS-CoV-2 Testing Consortium, Dirk Hockemeyer, Fyodor Urnov, Ralph Green, Jennifer A. Doudna

**Affiliations:** University of California, Berkeley, Berkeley, CA, USA; Innovative Genomics Institute, University of California Berkeley, Berkeley, CA, USA; UC Davis School of Medicine, Sacramento, California, USA; Howard Hughes Medical Institute, University of California, Berkeley, CA, USA; Gladstone Institutes, University of California, San Francisco, CA, USA

## Abstract

The appearance and spread of the novel severe acute respiratory syndrome coronavirus (SARS-CoV-2) led to the official declaration of a global pandemic, with states in the US implementing shelter-in-place orders at an unprecedented scale. SARS-CoV-2 has a robust person-to-person transmission rate and an asymptomatic period of two weeks or more, leading to widespread infection that has overwhelmed healthcare infrastructures around the globe. Effective public health measures require extensive, accurate, and rapid testing to determine infection rates. Here we describe the strategy we used to establish a CLIA-licensed clinical laboratory to perform a validated Laboratory-Developed Test (LDT) for SARS-CoV-2 in Berkeley, California and the surrounding Bay Area community. Our procedures for implementing the technical, regulatory, and data management workstreams necessary for clinical sample processing provide a roadmap to aid others in setting up similar testing centers.

**Note on Nomenclature:** *in accordance with established virology and infectious disease nomenclature, throughout this document we use “SARS-CoV-2” to refer to the viral agent causing infection and “COVID-19” to refer to the human infectious disease caused by that viral agent*.

## Introduction

The World Health Organization (WHO) declared a global pandemic on March 11, 2020 due to the spread of SARS-CoV-2 infections worldwide^1^. SARS-CoV-2 causes the coronavirus disease of 2019, or COVID-19, and continues to spread globally. As of April 6, over1,346,299 individuals are known to be infected, and more than 74,744 deaths have occurred^2^. At the time of this manuscript submission, there are no clinically validated medical interventions to prevent or cure COVID-19 cases. The public health focus has been on mitigating spread through diagnostic testing, self-isolation, and shelter-in-place orders in the US and elsewhere^3^.

While the majority of COVID-19 cases are asymptomatic or mild (80% according to one study^4^), a subset of patients can develop severe pneumonia leading to a reported mortality of ∼2%^5,6^. Countries such as Iceland, where extensive population testing is being performed, have reported that the age-wise distribution for the infected population is skewed toward a younger age range that largely presents with mild to no symptoms^7^. This indicates that the presence of these carrier and mildly symptomatic individuals in the general population may be a central driver for the outbreak. Notably, SARS-CoV-2 has a robust person-to-person transmission pattern, leading to the accelerated and widespread outbreaks that have overwhelmed the healthcare system infrastructure, causing additional deaths^3^. Given the lack of clinically validated treatment options, high transmission rates, and a large asymptomatic or mildly presenting patient population, community testing is central to controlling SARS-CoV-2 spread by quickly identifying, isolating, and treating infected patients and limiting transmission; this approach has been adopted by several countries, most notably Germany, and has resulted in lower mortality rates from COVID-19 compared to most other countries^8,9^. The Innovative Genomics Institute (IGI) at the University of California, Berkeley (UC Berkeley) addressed this challenge by establishing a clinical testing laboratory on a campus without a medical school in a three-week process described here.

Since the start of our effort, on March 14, 2020, the availability and turnaround time of SARS-CoV-2 testing at UC Berkeley and in the City of Berkeley have been inadequate relative to the public health need. Specifically, the turnaround time for testing for UC Berkeley students exceeded seven days (UC Berkeley Tang Center, personal communication). In addition, no rapid turnaround testing was available to City of Berkeley first responders, jeopardizing operational readiness. For example, multiple firefighters had to be quarantined following the diagnosis of COVID-19 in one firefighter (City of Berkeley Fire Department Chief David Brannigan, personal communication). Furthermore, no rapid testing or surveillance testing was available for vulnerable populations in Berkeley, including individuals who live in communal housing and the homeless.

Three regulatory developments allowed the IGI to rapidly transition its research laboratory space into a clinical testing facility. The first was the FDA’s March 16th Policy for Diagnostic Tests for Coronavirus Disease-2019 during the Public Health Emergency (see *Useful Links*). This policy simplified the process for getting authorization for a testing method and workstream. The second was California Governor Newsom’s Executive Order N-25-20 (see *Useful Links*), which modified the requirements for clinical laboratory personnel running diagnostic tests for SARS-CoV-2 in a certified laboratory. The third was increased flexibility and expediency at the state and federal levels for certification and licensure requirements for clinical laboratory facilities under the Clinical Laboratory Improvement Amendments (CLIA) program. Under these emergency conditions, the California Department of Public Health (CDPH) was willing to temporarily extend, once the appropriate regulatory requirements have been fulfilled, an existing CLIA certificate for high-complexity testing to a non-contiguous building on our university campus. These developments enabled us to develop and validate a Laboratory-Developed Test (LDT) for SARS-CoV-2, extend the UC Berkeley Student Health Center’s clinical laboratory license to our laboratory space, and begin testing patient samples.

Our process comprised:

1. partnering with the UC Berkeley Health Services Center and certified Clinical Laboratory;
2. notifying Centers for Medicare & Medicaid Services of the intention to extend the CLIA certificate;
3. selecting an FDA-approved commercial testing platform as the basis for establishing our LDT, and validating the resulting LDT in accordance with CLIA guidelines;
4. training and registering graduate student and postdoctoral volunteers with the California Department of Public Health (CDPH) in place of clinical laboratory personnel to run the tests;
5. establishing a custom Laboratory Information Management System (LIMS) for Health Insurance Portability and Accountability Act (HIPAA)-compliant resulting;
6. obtaining a Biological Use Authorization from UC Berkeley’s institutional biosafety committee (the Committee for Laboratory and Environmental Biosafety, CLEB), involving determining the level of biosafety (BSL-2+) and nature of personal protection equipment (PPE) necessary to accession the patient samples and process them;
7. ensuring all testing personnel obtain a certificate of HIPAA training to handle Personal Health Information (PHI);
8. establishing an interface to University Health Services (UHS) and LifeLong Medical Care, which serves vulnerable populations (e.g. the uninsured and the homeless), that allows both test requisition in our diagnostic facility and sample resulting to the physician who requisitioned the test in a HIPAA-compliant fashion, as well as reporting the testing summaries to the California Reportable Disease Information Exchange (CalREDIE) of the State of California Department of Public Health;
9. obtaining a temporary extension to our facility of the CLIA certificate held by the UC Berkeley Health Services Laboratory for high-complexity testing; and
10. submitting an Emergency Use Authorization (EUA) request for our LDT to the FDA.

To achieve this formidable task, the IGI team divided into specialized subteams responsible for establishing, and then executing in parallel, all necessary procedures on the technical, supply chain, regulatory, human resources, data management, sample collection, and sample reporting processes necessary for laboratory operations. We initiated the effort on March 16, 2020 and processed our first patient samples using our LDT three weeks later, on April 6, 2020. Thus, with a dedicated and structured three-week effort, this team developed, validated, and operationalized the LDT described here in a CLIA-compliant manner, enabling SARS-CoV-2 PCR-based testing and reporting of results from patient samples in collaboration with our clinical partners.

Here, we provide a comprehensive description of our IGI SARS-CoV-2 Diagnostic Testing Laboratory, documenting our strategy for addressing all relevant regulatory and infrastructure requirements, which allowed us to enable community testing in the Berkeley area in our IGI/UC Berkeley UHS Diagnostic Testing Laboratory.

## Results

### ESTABLISHMENT OF A CLINICAL TESTING FACILITY

#### Ensuring Regulatory Compliance, Part 1: Clinical Laboratory Improvement Amendments Certification Process

##### Clinical Laboratory Improvement Amendments (CLIA) program

The Centers for Medicare & Medicaid Services (CMS) regulate all clinical laboratory testing performed in the U.S. through the [Clinical Laboratory Improvement Amendments (CLIA)](https://www.cms.gov/Regulations-and-Guidance/Legislation/CLIA/index?redirect=/clia/) program. CLIA was established to ensure accuracy and reliability of patient test results. There are certification and licensure requirements for both the facility and the staff, which are based on the types of tests performed. The federal CLIA program contracts through State Agencies like state public health departments for oversight and implementation of the program (See *Useful Links* for State Survey Agency CLIA contacts).

Tests are categorized as waived, moderate complexity, or high complexity based on the test’s “score” across seven categories considered for CLIA test categorization (see *Useful Links* for CLIA Test Categorization). A SARS-CoV-2 qPCR-based test is considered “high complexity”. For a high-complexity test, the CLIA certified laboratory must hold a special certificate ensuring its ability to perform these more complex tests (“Certificate of Compliance,” surveyed by CMS, or a “Certificate of Accreditation,” surveyed by private accrediting organizations).

Typically, for CLIA certification, clinical laboratories must apply for and receive both a state license and a federal CLIA certificate, usually a lengthy process. In this emergency pandemic situation, temporarily relaxed regulatory requirements and expedited review by CMS and the California Department of Public Health made it possible to extend an existing CLIA certification and state license for high-complexity testing from a CLIA-certified clinical laboratory on our campus, UC Berkeley’s UHS clinical laboratory, to temporarily include the testing facility at the IGI. This secondary site will operate for the duration of the State of Emergency, as defined by the State of California and the federal government.

##### CLIA Personnel Qualifications

CMS and state law dictate specific requirements for personnel in CLIA laboratories that are particular to the category of testing being run. California state law requires that clinical laboratories employ testing personnel who are licensed by the California Department of Public Health. However, following the COVID-19 outbreak in the US, on March 4, 2020, the Governor of California issued Executive Order N-25-20, which temporarily suspended the requirement for licensure of personnel performing testing for SARS-CoV-2, so long as the personnel meet the requirements under section 353 of the Public Health Service Act, and conduct the testing in a facility certified and licensed to perform high-complexity testing. The Branch Chief of CDPH, Robert J. Thomas, further clarified in his March 24th letter to Laboratory Directors and Managers that while the executive order removes the licensing requirements for testing personnel, it does not alter the requirements for a laboratory director, clinical consultant, technical consultant, technical supervisor, and general supervisor. Thus, while the governor’s executive order allowed us to have UC Berkeley scientists who are not licensed Clinical Laboratory Scientists (CLS) to develop and implement our testing pipeline and to recruit scientist volunteers to act as testing personnel to run the samples, it preserved the essential requirement to have licensed CLSs help to set up and oversee our testing facility.

Thus, a close working relationship with the UHS Clinical Testing Laboratory was essential to our success, and UHS CLSs and its Laboratory Director provided key guidance regarding the development of our workflow and navigation of CLIA regulations. Further direct communication with CDPH Laboratory Field Services (LFS) led CDPH to approve extension of UC Berkeley’s existing CLIA certificate for high-complexity testing at UHS to include a temporary second site at the Innovative Genomics Institute Building, a UC Berkeley facility, for emergency SARS-CoV-2 testing. Extending this existing CLIA certificate allowed us to work under the supervision and guidance of the Tang Center Clinical Lab’s Laboratory Director and Pathologist, and with licensed CLSs in the laboratory,, meeting the requirements under Governor Newsom’s executive order. Their participation in our effort was critical from a compliance perspective and has been invaluable in the scientific development of our workflow.

##### Requirements for Training Under CLIA

Chief Thomas stated in his March 24th letter that, “the laboratory director is responsible for the competency assessment and documentation of all personnel testing for SARS-CoV-2. [and] The laboratory director must provide the list of testing personnel along with documentation of competency upon request by LFS.” We therefore developed and documented a training strategy for our UC Berkeley scientist volunteers under the guidance of the Tang Center’s CLS staff, and submitted evidence to CDPH LFS of prior training (education) in a relevant field by submitting their CVs and either transcripts, diplomas, or letters from PhD thesis advisors attesting that the relevant individuals possessed an advanced degree. While the specific educational requirements have been relaxed during this emergency public health crisis, CLIA regulations still require that all testing personnel be rigorously trained and evaluated, both initially and periodically, to ensure quality performance.

#### Ensuring Regulatory Compliance, Part 2: Personnel Safety and Privacy and Security of Protected Health Information

Beyond a CLIA license, the IGI testing facility required additional regulatory compliance to perform testing for SARS-CoV-2 in patient samples. The IGI testing facility obtained a Biological Use Authorization (BUA) from UC Berkeley’s institutional biosafety committee, the Committee for Laboratory and Environmental Safety (CLEB), which determined the level of biosafety and PPE required for accessioning and processing patient samples.

To ensure the safety of our staff, virus in patient samples is inactivated at the time of sample collection (see “*Sample Collection*” in Methods), meaning that infectious virus is never present in our testing facility. As a further precaution, our laboratory operates under BSL-2+ conditions, providing a higher standard of safety than the BSL-2 conditions typically employed with inactivated SARS-CoV-2. Especially stringent PPE requirements were established, including a disposable outer layer and N95 masks fitted for each staff member. The additional precautions in our level of biosafety and PPE result from our determination to maximally protect the health of our volunteer staff (see *Personnel Biosafety and HIPAA Training* section below).

Our sample collection kits are returned to the testing facility by our health partners containing patient-identifying information, and thus HIPAA compliance is observed in our laboratory procedures and by our personnel. To ensure HIPAA compliance, the laboratory space cannot be accessed physically or visually by anyone other than trained personnel. Laboratory windows have closed blinds to ensure no visibility to the inside of the lab is possible, and no photography or video is permitted in the laboratory space. Additionally, no patient health information can be shared outside of a HIPAA-compliant communication method, and therefore our laboratory information management system (LIMS) is built ensuring HIPAA compliance both in the semi-manual and automated approaches of our testing workflow (see below under the *Laboratory-Developed Test* Section). All data that are generated by instruments in the laboratory are isolated from external networks, and instrument data are transmitted to the LIMS using secure protocols. Additionally, instrument data are periodically purged from laboratory instruments and stored on an encrypted storage server in a hardened secure location. Moreover, any person entering the lab must have completed HIPAA training (see *Personnel Biosafety and HIPAA Training* below).

##### Personnel Biosafety and HIPAA Training

All testing laboratory staff members are trained in best practices related to BSL-2 according to our BUA, HIPAA, PPE, as well as general laboratory procedures and safety, prior to starting laboratory testing work. HIPAA training was provided by the Thomson Reuters HIPAA Privacy and Security (California) course, which covers fundamental HIPAA privacy and security principles, such as administrative, physical, and technical safeguards, and handling and use of protected health information. An additional chapter of the course covers medical privacy laws of California, which include requirements concerning the handling and use of protected health information and penalties for violations under California state law. Personnel involved in accepting, accessioning, testing, and resulting the patient samples were required to take the class and pass the exit examination before beginning work in the facility. Non-testing staff who will be present in a laboratory in which patient health information exists must also be HIPAA-trained (e.g. equipment technicians).

The supplementary file, *Personnel Training and Compliance*, provides details on laboratory and biosafety procedures, as well as the full training checklist required from each laboratory staff member before they can commence work.

To minimize the risk of infectious individuals entering the building where the diagnostic laboratory is housed, 2-4 hours prior to entry the personnel complete a self-screening questionnaire (see supplementary file, *Personnel Training and Compliance* for our Daily Self Assessment Questionnaire). When entering and moving within the testing facility, all individuals, including equipment technicians and supervisors, must wear appropriate PPE at all times. To enter the testing facility, personnel must put on PPE in the following order: lab coat, disposable isolation gown tied in the back, double gloves with inner gloves over cuffs, N95 respirators, and face shields. Additionally, ankles must not be exposed and all personnel must wear closed shoes, preferably a specific pair that is worn only in the testing laboratory. Disposable PPE items (gowns, gloves) do not leave the testing facility unless enclosed in a biohazard bag that is sprayed externally with 70% ethanol. N95 respirators must be rigorously fitted to each staff member. This fitting is performed by a UC Berkeley EH&S expert who verifies air flow and sealing to ensure protection of the user. For more details on PPE requirements for our personnel, see supplementary file, *Personnel Training and Compliance*.

### ASSAY DEVELOPMENT

#### Laboratory-Developed Test (LDT)

Several commercially-available diagnostic tests for detecting SARS-CoV-2 viral RNA using reverse transcription combined with the polymerase chain reaction (RT-PCR) have received an EUA from the FDA. The IGI chose the test developed and marketed by Thermo Fisher Scientific (Fig. 1A, B) due to availability of test reagents, compatibility with our equipment, and the robust test performance in our hands in pilot experiments. However, because we enacted modifications to the EUA workflow, our implementation of the Thermo Fisher test became a laboratory-developed test (LDT) as defined by CMS and the FDA^10^. We therefore had to establish analytical and clinical validity of our adaptation of the test through experiments determining our limit of detection, and through bridging assays comparing our results to those of other authorized testing sites.

**Figure 1.**
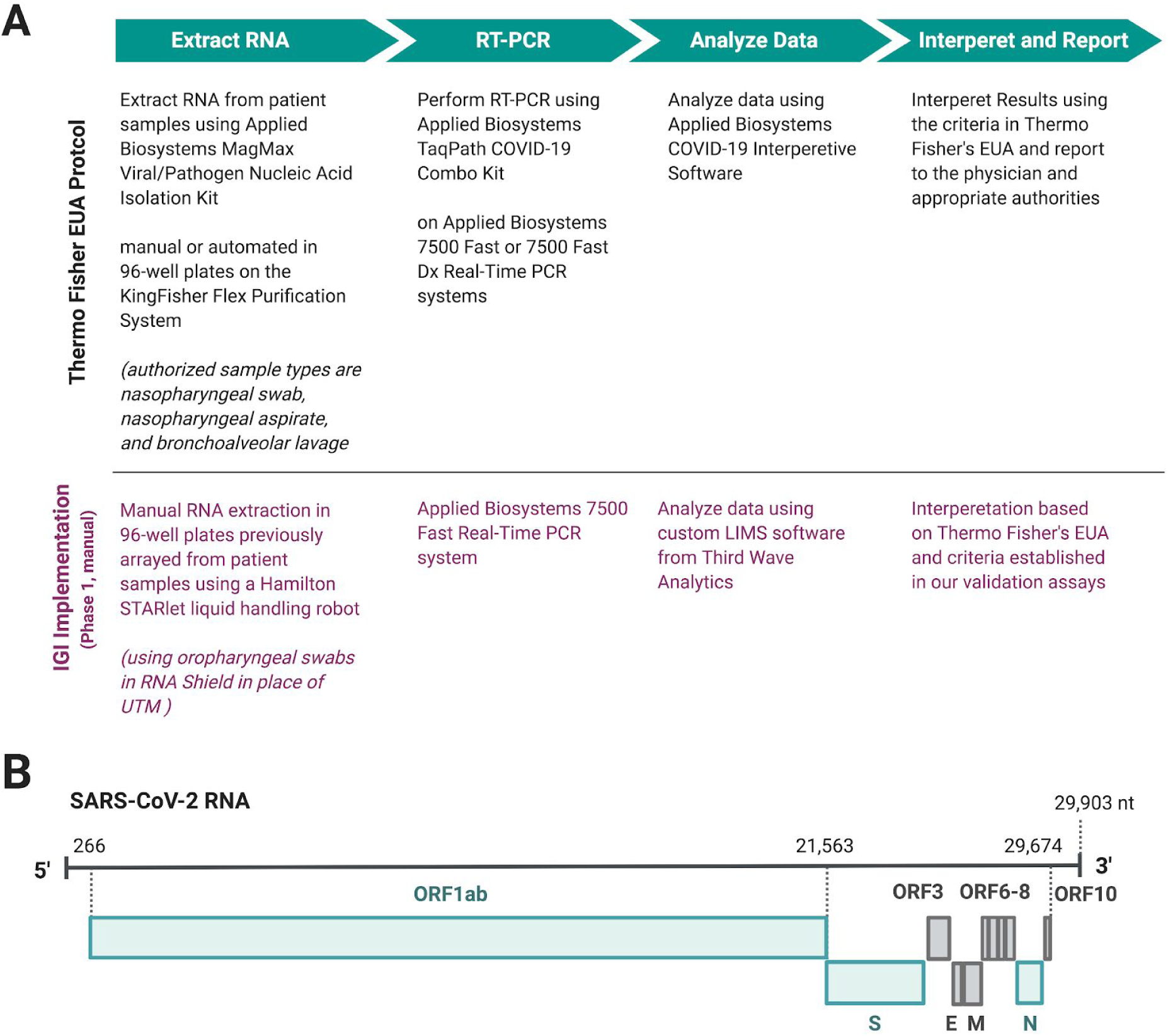
Overview of the Innovative Genomics Institute SARS-CoV-2 Testing Consortium (IGI Consortium) assay and those discussed in this report. **A**: Overview of Thermo Fisher’s TaqPath COVID-19 Multiplex Diagnostic Solution^11,12^ showing the steps as authorized in their EUA (black), and IGI’s manual implementation of this workflow (magenta). The IGI’s SARS-CoV-2 testing lab was established in two phases. Phase 1 started with automated arraying of patient samples, followed by manual implementation of the Thermo Fisher kit. Phase 2 is a fully automated workflow. **B**: SARS-CoV-2 genome (NCBI ref NC_045512.2) with translated regions diagrammed in rectangles. Genes targeted by Thermo Fisher’s kit are shown in teal^11^, while other open reading frames and gene products are shown in grey. The open reading frame, ORF1ab, targeted by Thermo Fisher’s kit encodes non-structural proteins for replication, while the spike (S) and nucleocapsid (N) genes encode two structural proteins. Figure created using BioRender.com.

Our decision to make modifications to the Thermo Fisher workstream was rooted in a desire to increase testing throughput, reduce cost, ensure safety of our volunteers, and make use of existing and donated equipment. To increase throughput, we are implementing multiple automated liquid handlers and using qPCR machines with increased capacity. To reduce cost and to protect against supply chain pressure, we tested and validated the use of Thermo Fisher’s RNA extraction and qPCR protocols at half the suggested reaction volumes (discussed in *Reduced Reaction Volumes* further on). To ensure the safety of our volunteers, we substituted the standard patient sample universal transport medium (UTM) for a virus-inactivating buffer (discussed in the *Sample Collection* section). With these modifications to the Thermo Fisher protocol, our LDT was developed and validated within the CLIA framework, and notice of the validation will be submitted to the FDA as a bridging study to the original EUA awarded to Thermo Fisher within the 15 day window specified by law.

Due to the complexity of establishing a fully automated workflow, and in the interest of supporting community testing efforts at the earliest possible time, we designed our workflow to support two parallel workstreams: a semi-manual approach and an automated approach. We initiated patient testing with the semi-manual approach at a capacity of ∼200 tests/day, while finalizing implementation of the automated approach for a capacity of 1000+ tests/day (see Table 1 for equipment used and differences between the two approaches; see Supplementary Figure 1 for the semi-manual approach workflow).

**Table 1.**
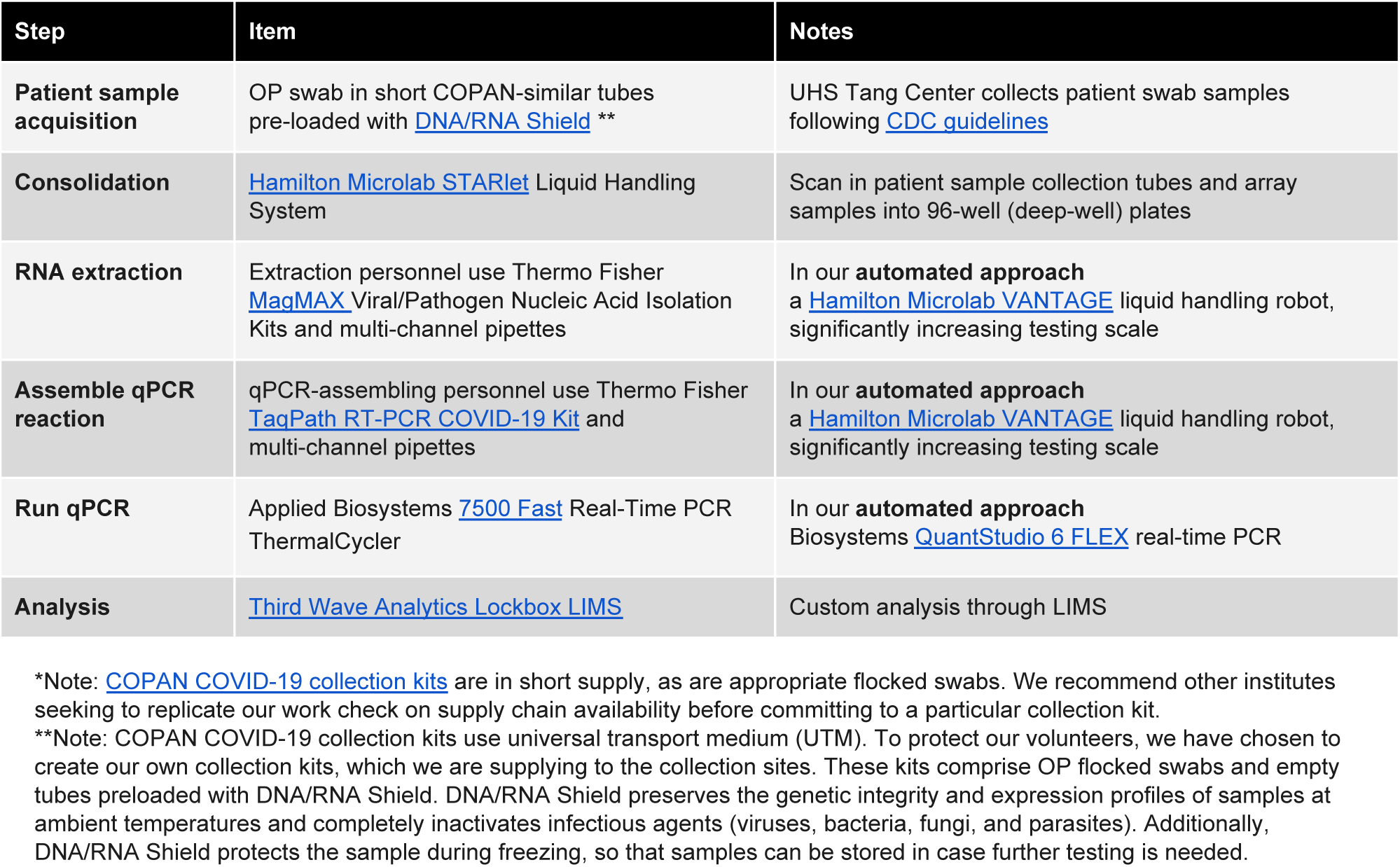
Critical equipment and reagents for IGI’s semi-manual workflow. The steps for RNA extraction, assembling the qPCR reaction, and running the qPCR reaction differ from the automated workflow, as described in the “Notes” column.

To automate workflows, integrate instruments, and manage patient sample data, we implemented a laboratory information management system (LIMS) that we customized to our processes and operational needs. The IGI testing facility uses [Third Wave Analytics Lockbox LIMS](https://thirdwaveanalytics.com/). Personnel from Third Wave Analytics customized the Lockbox LIMS based on the needs of both our semi-manual and automated approaches in a cost-effective and HIPAA-compliant way, with the intent of implementing the system in two main phases. Phase one of the build, corresponding to our semi-manual approach, established the core laboratory operating system to manage de-identified barcoded sample data and operational data as testing occurs, including highly automated sample record creation, testing data management, and data auto-analysis. The final output of the Lockbox LIMS is an analyzed sample result, awaiting approval for final reporting by appropriate laboratory personnel.

For phase one, to ensure HIPAA compliance in the semi-manual LIMS, no PHI is entered into the LIMS, and only de-identified patient barcodes are used for sample tracking. Upon reporting by our health partners at the UHS Tang Center, the LIMS returns a comma separated value (csv) file including de-identified barcoded results. These are sent to the Tang Center for integration in their electronic health system, where the patient results are connected to a specific patient’s file via barcode matching. In phase two of the LIMS build, corresponding to our fully automated approach, we have implemented a HIPAA-compliant portal through which tests will be requested and reported to enable testing for non-UC Berkeley patient samples, given the large-scale capacity of the fully automated LDT operation (1000+ tests/day).

Third Wave Analytics partners with institutions looking to establish SARS-CoV-2-related clinical testing, and knowledge gained from our partnership can be leveraged to rapidly develop and implement an appropriate LIMS for a similar custom setup. For more details on the LIMS interface with our workflow, please see Supplementary Figure 1 and supplementary video, *Semi-Manual LIMS Interface*.

#### Establishment of a Novel Sample Collection Kit

The standard sample collection kits use COPAN or similar tubes with universal transport medium (UTM) and nasopharyngeal (NP) or oropharyngeal (OP) swabs. NP swabs are preferred because of the higher viral load in collected samples, though both types are accepted as per CDC guidelines. Due to bottlenecks in supply of NP swabs and standard COPAN tubes, we currently use OP swabs in our sample collection kit, as well as shorter COPAN-type tubes that we adapted to our setup by 3D-printing custom inserts to fit our Hamilton Microlab STARlet tube racks (see Supplementary Fig. 1 for details on our Hamilton Microlab STARlet setup for accessioning patient samples). UTM maintains the activity of the virus and stabilizes it during transport. To avoid receiving live virus at the testing site, other research centers transitioning to clinical testing sites have instituted RNA extraction at the medical facility prior to delivery to the testing facility. In our case, our partner health center did not have the capacity to perform pre-testing preparation. To protect our staff from active virus arriving on site, we created our own sample collection kits using DNA/RNA Shield from Zymo Research instead of UTM. These kits are delivered to sample collection sites (the first site being the Tang Center on the UC Berkeley campus). DNA/RNA Shield inactivates viruses and stabilizes nucleic acids for transport at room temperature.

##### Interfacing with Clinicians

The IGI is working to develop relationships with multiple health care providers, and each partnership requires development of a workflow that works for both parties. For instance, since we developed an LDT under the Tang Center license, we have worked closely together with Tang Center clinicians and clinical laboratory to establish how specimen collection and processing occurs for their student population. Separately, the City of Berkeley Public Health Department has established a partnership with LifeLong Medical Care to requisition tests and collect specimens for first responders (e.g. firefighters) and the homeless. The IGI is partnering with LifeLong and receiving samples from these individuals as well. One challenge has been to design sample collection kits and clinician protocols for both groups, such that specimens arriving at the IGI from different sources can be accessioned and tested together. Integrating CLIA-grade best practices into all steps of the testing pipeline was an additional challenge: it is the responsibility of the IGI to ensure that resulting is accurate, and this requires building redundancy into the pipeline and checks at multiple steps.

Batches of SARS-CoV-2 collection kits are assembled as needed and quality-controlled at the IGI (see below). These kits are designed for ease of use easily by Tang Center and LifeLong clinicians and nurses. Tang Center and LifeLong have different electronic medical records (EMR) management systems, and have different mechanisms for connecting a patient record to a barcode that is compatible with our robotic accessioning step. As such, their kits differ in whether the IGI provides barcodes or whether the barcodes are generated within the healthcare system. The Tang Center and LifeLong are responsible for requisitioning a test for a patient, collecting the specimen, and integrating that sample into their EMR. Following transport to the IGI, the acquisition personnel perform multiple checks to confirm that all barcodes and patient identifiers, if present, match those on tubes, within kits, and on requisitions or manifests. Samples that do not match are rejected for specimen re-collection. With a verified barcode, the LIMS tracks specimen progress through the testing pipeline and returns a result. That result needs to be re-matched with a patient record when testing is complete and reported.

### ASSAY VALIDATION

#### Reduced Reaction Volumes

To lower the overall cost of our assay per test, and to limit reagent use in the face of supply chain shortages, we sought to reduce the quantity of reagents used per RNA extraction and RT-qPCR assay. After pilot experiments demonstrated that the test performed robustly after such a reduction, we proceeded to validate our workflow at half-reaction sizes (see supplementary file, *Semi-Manual SOP*, for exact volumes). In the following section, we report the assay validation data, all conducted with the reduced volume protocol, which comprised our validation report to CDPH and the basis of our FDA EUA submission.

#### LDT Validation

Validating an LDT requires measuring specific FDA/CDC-defined metrics and meeting or exceeding benchmarks. In the case of our SARS-CoV-2 LDT, these metrics were: (i) measuring assay limit of detection (LOD); (ii) assessing clinical and analytical validity by running mock positive and negative samples at known concentrations, (iii) performing our LDT on samples from SARS-CoV-2-positive and -negative individuals provided by two local clinical diagnostic testing facilities. This allowed us to bridge results from our LDT to those attained by CLIA-certified diagnostic laboratories performing distinct LDTs with existing or submitted EUAs.

##### Limit of Detection

The March 16th FDA guidance recommends identifying the LOD of a molecular diagnostic by conducting a dilution series of SARS-CoV-2 RNA (or inactivated virus) in an artificial or real clinical matrix. The FDA recommends that “laboratories test a dilution series of three replicates per concentration, and then confirm the final concentration with 20 replicates.” For the purposes of an EUA, the FDA defines the limit of detection as “the lowest concentration at which 19/20 replicates are positive.” In this section we describe the experimental data that establish the LOD of our LDT.

To experimentally determine our limit of detection, we performed serial dilutions of SARS-CoV-2 positive control RNA (see Methods) in triplicate, from 5 × 10^4^ to 1 × 10^2^ copies/ml in 500 copies/ml steps. At each dilution, we determined the Ct (cycle threshold) of three SARS-CoV-2 genes (N-gene, S-gene and ORF1ab) as well as spiked-in MS2 bacteriophage nucleic acid, an internal control for nucleic acid extraction efficiency and qPCR amplification. The Ct value is inversely proportional to the amount of starting target RNA, and represents the PCR cycle number at which the fluorescent signal of the reaction crosses a set threshold. This threshold was set manually based on background fluorescence of the reaction mixture.

Across the six dilutions tested, we observed a mean Ct value for MS2 of 24, with little variability among samples, indicating consistent nucleic acid RNA extraction (Fig. 2A). At our lowest concentration tested, 1 × 10^2^ copies/ml, we were not able to detect amplification of any of the three SARS-CoV-2 primer-probe sets in any of our replicate samples, despite consistent MS2 detection, indicating that 1 × 10^2^ copies/ml is below our detection limit (Fig. 2A, left). At 5 × 10^2^ we observed amplification of each of the three target SARS-CoV-2 genes, however these values were not reproducible across triplicates (Fig. 2A, ORF1ab is undetected for a single replicate at 5 × 10^2^ copies/ml). Thus, the limit of detection was identified as 1 × 10^3^ copies/ml, the lowest concentration tested where all target genes were detected for all replicates. Extraction and PCR controls show expected values (Fig. 2B), with SARS-CoV-2 Ct values exclusively detected in the PCR positive control and MS2 Ct values detected in the RNA extraction and PCR MS2 spike-in controls.

**Figure 2.**
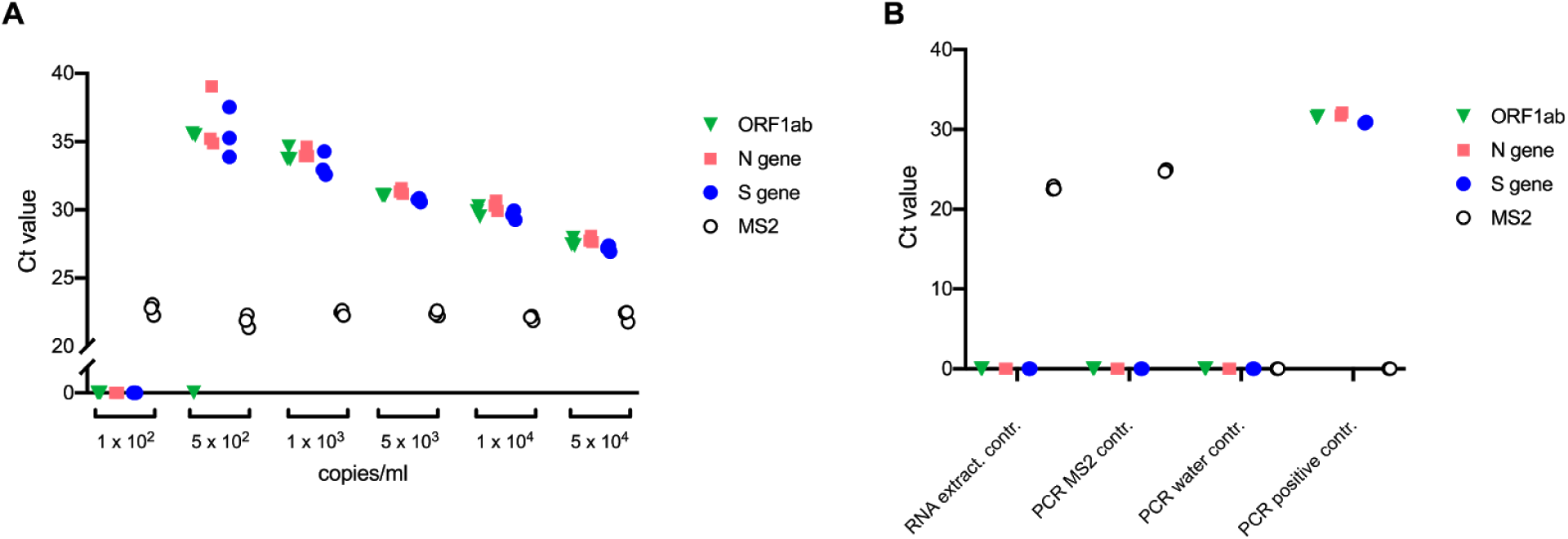
Limit of detection assay. **A**: Serial dilutions of SARS-CoV-2 positive control RNA. At each concentration, RNA extraction and subsequent RT-qPCR was performed in triplicate using primers and probes targeting ORF1ab, N gene, S gene, and MS2 RNA internal control. All replicates are plotted as individual points clustered by primer/probe targets. The concentration of 1 × 10^3^ copies/ml was the lowest dilution at which all replicates returned positive Ct values for all target genes. For clarity and resolution of data points, the y axis is plotted as a broken axis. An undetermined Ct value is plotted as Ct = 0. **B**: Quality controls. An RNA extraction control (N = 3) containing the kit’s MS2 control in sample collection buffer (DNA/RNA Shield in PBS) is used as an RNA extraction, reverse transcription, and PCR amplification control. Additional quality controls, in duplicate, are added during preparation of RT-qPCR plates (after RNA extraction) and include, from left to right, MS2 RNA in PCR master mix, water negative control in PCR master mix, and SARS-CoV-2 positive control RNA in PCR master mix. All replicates are plotted as individual points clustered by primer/probe targets. An undetermined Ct value is plotted as Ct = 0.

We next sought to confirm the measurement 1 × 10^3^ genomic copies/ml as the LOD by performing the assay in 20 technical replicates. For each of the 20 replicates, we detect all three SARS-CoV-2 genes, as well as MS2 internal control, as demonstrated by consistent, positive Ct values (Fig. 3, left). To control for non-specific amplification, we extracted human RNA from 293T cells and subjected it to the same RNA extraction and RT-qPCR workflow. We observed no cross reactivity from any of the four primer-probe sets (Fig. 3, full workflow controls). Nucleic acid extraction and PCR controls showed expected values, with no detectable amplification of SARS-CoV-2 genes in MS2 nucleic acid only or water only samples (Fig. 3, PCR controls). The SARS-CoV-2 positive control from Thermo Fisher exhibited expected amplification of all three SARS-CoV-2 genes targeted, as well as the expected absence of MS2 nucleic acid. Together, our results show that we were able to detect 20/20 of our replicates, exceeding the required 19 out of 20 as defined by the FDA, and establishing 1 × 10^3^ genomic copies/ml as our LoD.

**Figure 3.**
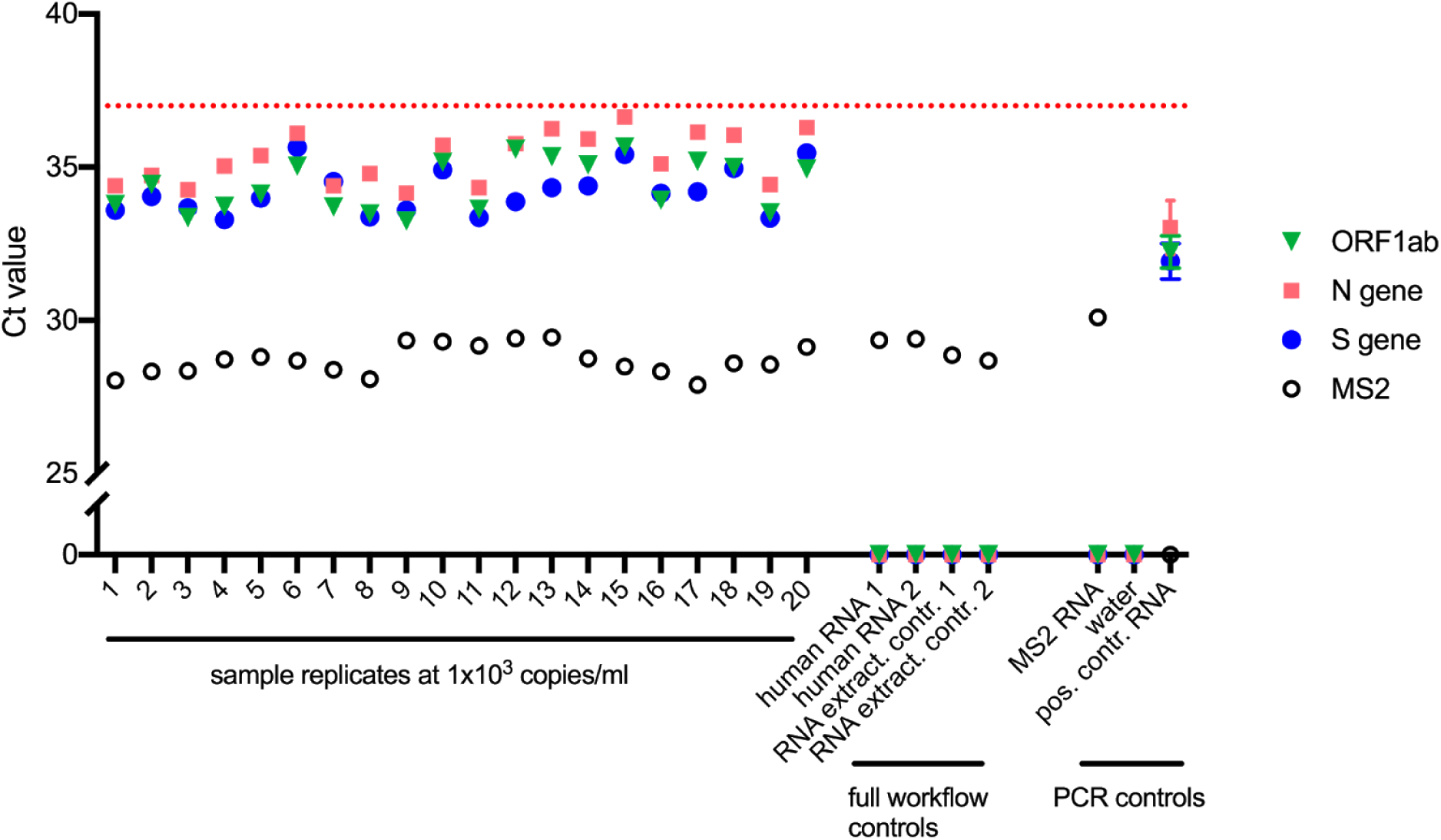
Confirmation of 1 × 10^3^ genomic copies/ml as the LoD. PCR controls (on the right) were performed in duplicate and are plotted as mean values with error bars representing standard deviation (SD). Values for individual replicates are plotted for all other samples. The y axis is displayed as a broken axis to allow better resolution at higher Ct values while still displaying Ct values of 0. An undetermined Ct value is plotted as Ct = 0. The red dotted line at a Ct of 37 indicates the maximum Ct value that will return a valid result as defined by the uppermost Ct values in LoD sample replicates.

##### Test Result Interpretation

At the concentration of our limit of detection, our highest Ct value was 36.67 (Fig. 3, sample 15, N gene). This defines an upper Ct boundary for reliably calling positive samples. From this, we set our reporting criteria such that a Ct above 37 will return a negative (NEG) result as defined in our analysis software for that gene.

For each sample, three SARS-CoV-2 genomic targets are assayed. To return a positive diagnosis, the EUA for Thermo Fisher’s TaqPath COVID-19 Combo Kit dictates that two out of the three gene targets must return a positive RT-qPCR result. In our setup this translates to a detected Ct value of less than 37 (but greater than zero) for two out of the three genes. The successful amplification of SARS-CoV-2 genomic targets can result in reduced MS2 amplification efficiency. Because of competition for PCR master mix reagents, it is possible that a highly amplified SARS-CoV-2 sample will present a low or undetected MS2 control. Thus, our decision criteria allow for a negative MS2 internal control when two out of three SARS-CoV-2 genomic targets test positive. To be able to return a negative result, the MS2 internal control must be positive. Additional criteria are listed in Table 2.

**Table 2.**
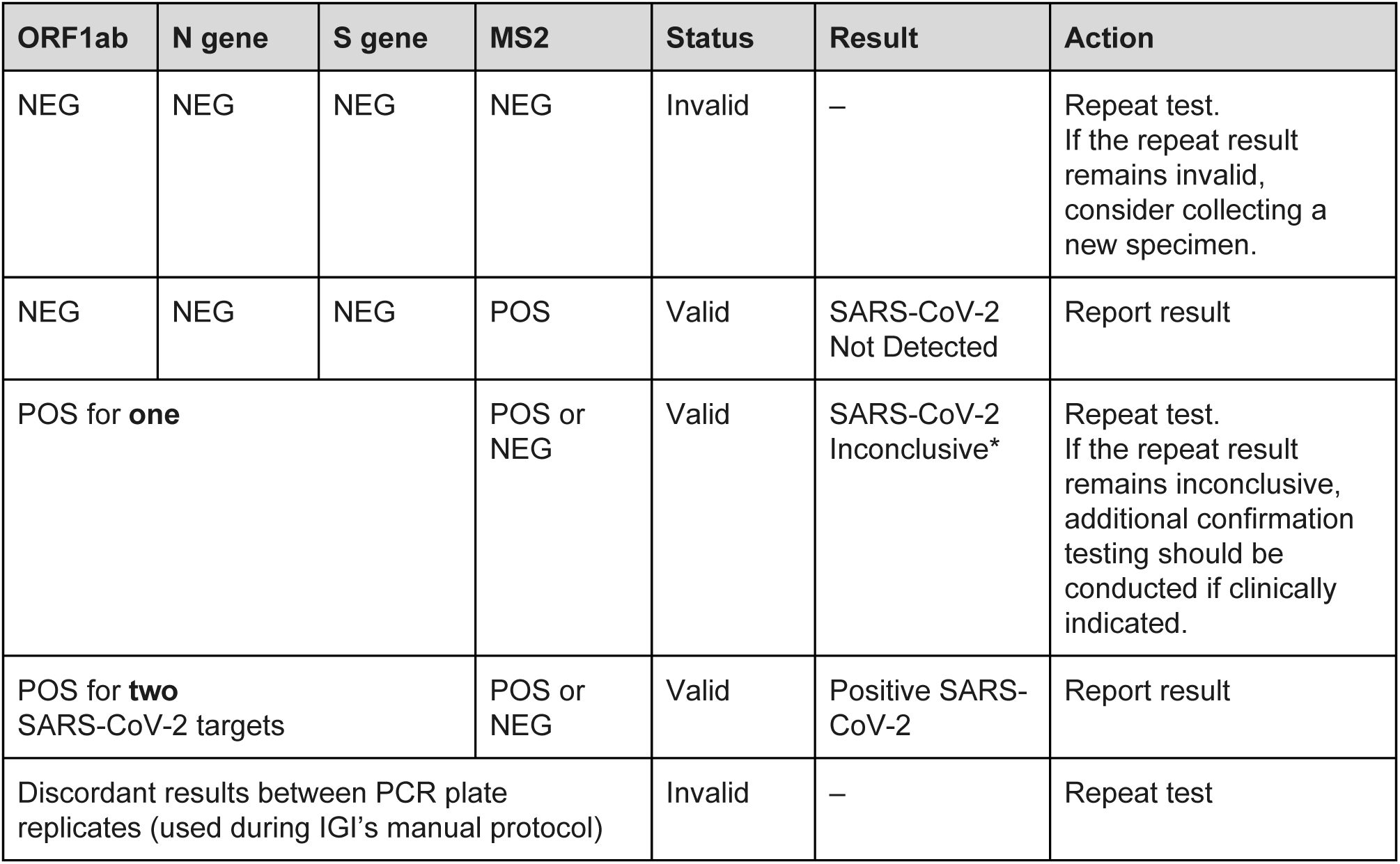
Result criteria and interpretation for patient samples. Reproduced and modified with permission from Thermo Fisher TaqPath COVID-19 Combo Kit Instructions for Use^11^. During our first phase of testing, we are using manual procedures for RNA extraction and qRT-PCR setup. As a manual pipetting control, we create duplicate PCR plates from the RNA extraction plate. The table above adds this outcome to the decision matrix to be used during our manual protocol only. \**Samples with a result of SARS-CoV-2 Inconclusive shall be retested one time*.

| ORF1ab | N gene | S gene | MS2 | Status | Result | Action |
| --- | --- | --- | --- | --- | --- | --- |
| NEG | NEG | NEG | NEG | Invalid | – | Repeat test.<br>If the repeat result remains invalid, consider collecting a new specimen. |
| NEG | NEG | NEG | POS | Valid | SARS-CoV-2 Not Detected | Report result |
| POS for **one** SARS-CoV-2 target |  |  | POS or NEG | Valid | SARS-CoV-2 Inconclusive* | Repeat test.<br>If the repeat result remains inconclusive, additional confirmation testing should be conducted if clinically indicated. |
| POS for **two** or more SARS-CoV-2 targets |  |  | POS or NEG | Valid | Positive SARS-CoV-2 | Report result |
| Discordant results between PCR plate replicates (used during IGI's manual protocol) |  |  |  | Invalid | – | Repeat test |

##### Clinical Sample Evaluation Assay

To ensure that diagnostic tests are clinically valid, the FDA recommends “that laboratories confirm performance of their assay with a series of contrived clinical specimens by testing a minimum of 30 contrived reactive specimens and 30 non-reactive specimens.” These reactive specimens can be created by spiking known concentrations of SARS-CoV-2 RNA or inactivated virus into leftover clinical specimens such as NP swabs in UTM. We chose to assess the clinical validity of our LDT by creating a range of different types of contrived SARS-CoV-2 RNA-positive and -negative samples using SARS-CoV-2 positive control RNA from the TaqPath COVID-19 Control Kit, SARS-CoV-2 patient samples from two local testing facilities, and human negative control RNA.

Our first experiment included 24 mock positive samples created by diluting Thermo Fisher’s SARS-CoV-2 positive control RNA from the TaqPath COVID-19 Control Kit into our sample collection medium (1X DNA/RNA Shield in PBS). The dilution range was chosen to span the LoD we had established above (Fig. 2). Additionally, 27 negative samples were created by diluting human RNA purified from 293T tissue culture cells. In the same plate, we included known positive and negative samples taken from leftover samples processed by a local clinical testing facility. These samples were provided to us in the sample collection medium that was used at the facility (universal transport medium) diluted 1:1 with 2x DNA/RNA Shield. To prepare these samples for our workflow, we further diluted them in our sample collection medium to 450uL before performing RNA extraction (see Methods). Figure 4 shows the results of this first clinical validation test. We have designed our workflow so that after RNA extraction, two replicate RT-qPCR plates are generated and run in parallel. Each plate must return concordant results to pass the criteria for interpreting positive or negative results. Results from one plate are shown in Figure 4 and the second plate results are in the supplementary material (Supplementary Fig. 2).

**Figure 4.**
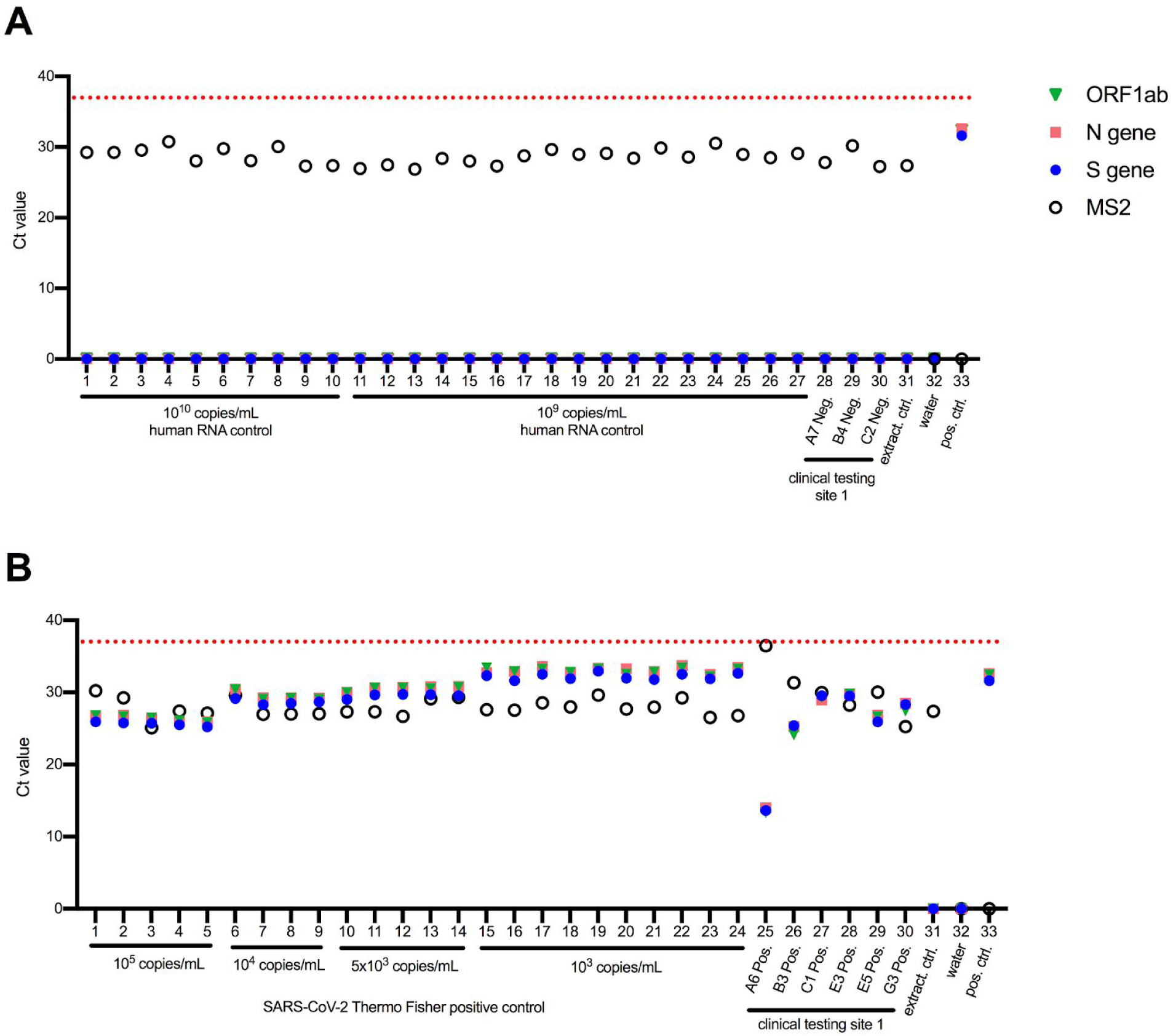
Clinical Validation Assay, Contrived and Clinical Samples - one of duplicate PCR plates. During our manual protocol we created duplicate PCR plates to control for human error. One of the two plates is shown here, the other can be found in Supplementary Figure 2. Replicate samples are plotted as individual points (rather than mean values). An undetermined Ct value is plotted as Ct = 0. **A**: Contrived negative and known negative samples from another local clinical testing site (clinical testing site 1). Whole cell RNA purified from cultured human 293T cells was diluted in IGI’s sample collection buffer (1X DNA/RNA Shield in PBS) to approximate negative samples. Human RNA concentration was calculated using UV absorbance, and the concentration was converted to an approximate copy number for comparison with positive RNA controls (See methods for details). Assay quality controls are shown in lanes 31–33, representing the extraction control (Thermo Fisher’s MS2 control in 1X DNA/RNA shield and PBS), and two PCR controls - water, and the Thermo Fisher kit positive SARS-CoV-2 RNA control (without MS2 nucleic acid). **B**: Contrived positive and known positive samples from another clinical testing site (clinical testing site 1). Contrived positive samples were prepared by diluting SARS-CoV-2 positive control RNA from Thermo Fisher’s kit into IGI’s sample collection buffer (1X DNA/RNA Shield in PBS). Assay quality controls are the same as in A.

To meet the clinical validity criterion, 95% of all SARS-CoV-2 RNA or virus-spiked samples must be positive, and 100% of all unspiked samples must be negative. As shown in Figure 4, our LDT attains the required sensitivity and specificity. We further validated our assay and workflow by testing additional known positive and known negative samples from a different testing facility. These samples (generous gift of Jeffrey Shapiro, Kaiser Permanente), were originally assayed at the Kaiser Permanente CLIA laboratory using the Roche cobas® SARS-CoV-2 kit targeting ORF1ab and E genes (Fig. 1) on a Roche Cobas8000 testing system, and were provided to us deidentified and bearing no PHI. Specimens for testing in our facility were created from the Kaiser samples by transferring the sample in UTM into a new tube with equal volume of 2X DNA/RNA Shield in PBS (our sample collection buffer) in a BSL-3 facility on campus. Once inactivated, decontaminated, and released from the BSL-3 lab, these samples were then run through our workflow at the IGI (Fig. 5 and Supplementary Fig. 3).

**Figure 5.**
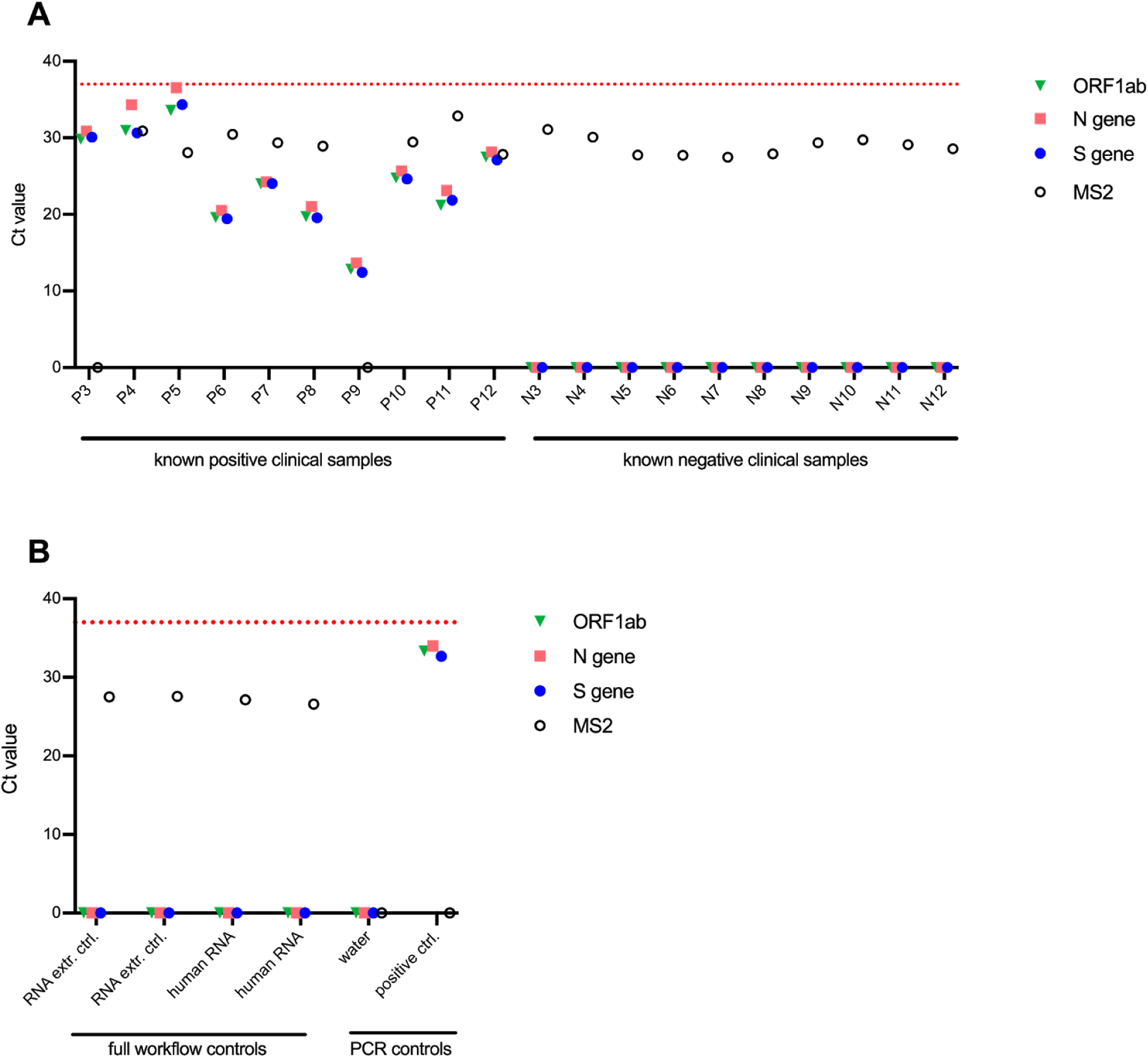
Clinical validation assay using leftover samples from Kaiser Permanente - one of duplicate PCR plates. The results from the second duplicate PCR plate are shown in Supplementary Figure 3. Undetermined Ct values are plotted as Ct = 0, and replicates are plotted individually. **A**: Ten known positive and ten known negative patient samples in UTM leftover from Kaiser Permanente’s clinical testing facility were delivered to Sarah Stanley’s BSL-3 lab at UC Berkeley where they were inactivated by diluting 1:1 in 2X DNA/RNA Shield. Decontaminated tubes were delivered to IGIB where they entered our testing pipeline, starting with arraying into 96-deep-well plates using the Hamilton STARlet robot. **B**: Assay quality controls. Full workflow controls were processed starting at RNA extraction in the 96-well plates. RNA extraction control is composed of Thermo Fisher’s MS2 control in 1X DNA/RNA shield and PBS. Human RNA controls are composed of 200 ng human RNA/ml in 1X DNA/RNA shield and PBS. PCR controls were processed starting at assembling RT-qPCR reactions and include a water control, and the Thermo Fisher kit positive SARS-CoV-2 RNA control (without MS2 RNA).

All 20 clinical samples returned results in concordance with the results reported by Kaiser Permanente (Table 3). Ct values for the known positive specimens spanned a wide range, providing good coverage of our dynamic range for clinical validation.

**Table 3.**
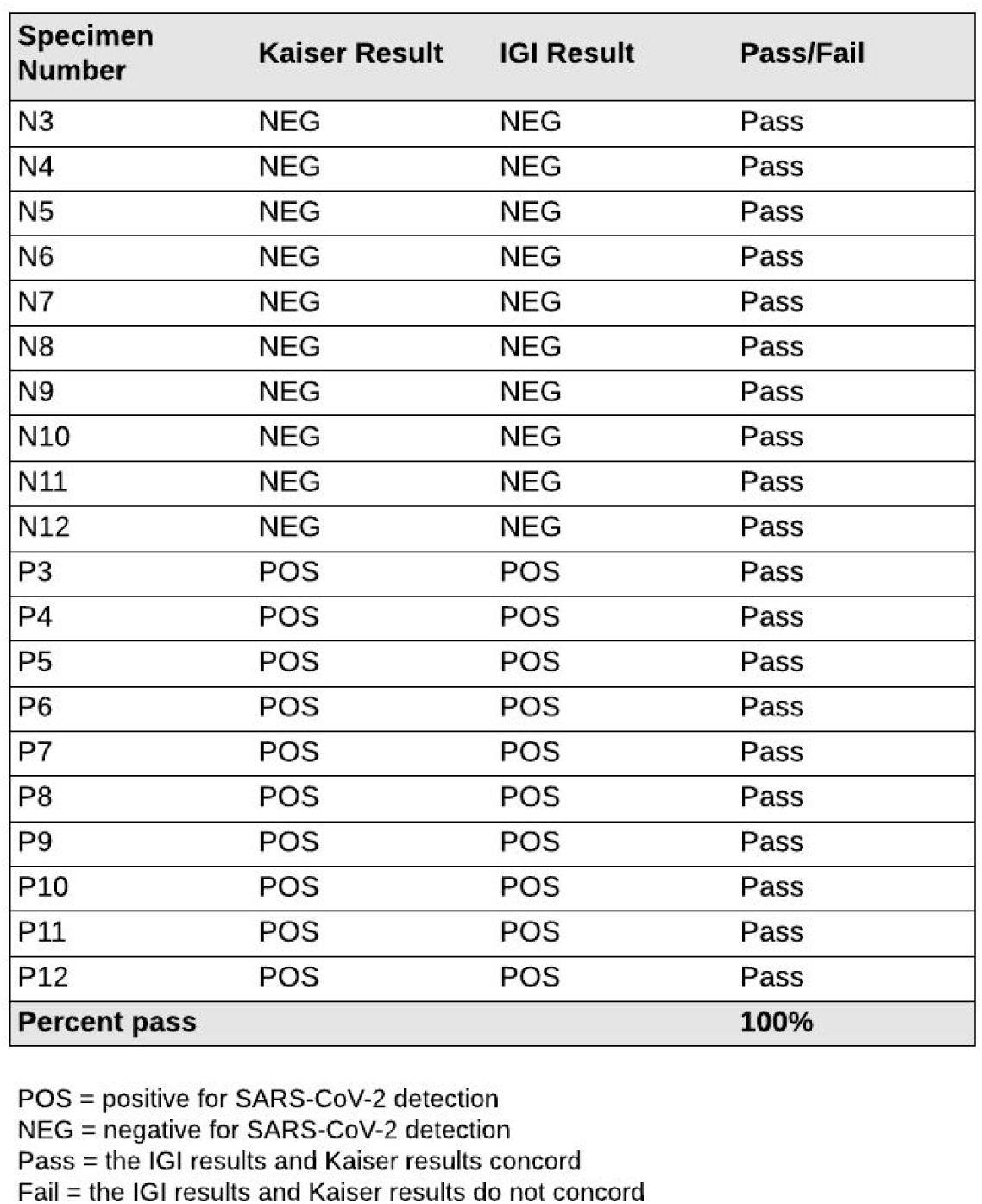
Concordance between the IGI’s assay results and external results for SARS-CoV-2 detection. 20 of Kaiser Permanente’s leftover clinical samples (10 positive and 10 negative) were run through the IGI’s SARS-CoV-2 detection assay. For the resulting decision matrix, see table 2.

| Specimen Number | Kaiser Result | IGI Result | Pass/Fail |
| --- | --- | --- | --- |
| N3 | NEG | NEG | Pass |
| N4 | NEG | NEG | Pass |
| N5 | NEG | NEG | Pass |
| N6 | NEG | NEG | Pass |
| N7 | NEG | NEG | Pass |
| N8 | NEG | NEG | Pass |
| N9 | NEG | NEG | Pass |
| N10 | NEG | NEG | Pass |
| N11 | NEG | NEG | Pass |
| N12 | NEG | NEG | Pass |
| P3 | POS | POS | Pass |
| P4 | POS | POS | Pass |
| P5 | POS | POS | Pass |
| P6 | POS | POS | Pass |
| P7 | POS | POS | Pass |
| P8 | POS | POS | Pass |
| P9 | POS | POS | Pass |
| P10 | POS | POS | Pass |
| P11 | POS | POS | Pass |
| P12 | POS | POS | Pass |
| **Percent pass** |  |  | **100%** |

POS = positive for SARS-CoV-2 detection
NEG = negative for SARS-CoV-2 detection
Pass = the IGI results and Kaiser results concord
Fail = the IGI results and Kaiser results do not concord

##### LIMS Verification

The previously tested and verified Lockbox LIMS software v1.48 was used as the template for the LIMS. Significant customizations were implemented to the v1.48 LIMS package, per the laboratory user requirements of the SARS-CoV-2 detection workflow (see *Methods* subsection *LIMS Customization* for an overview of all customizations).

Prior to clinical sample testing, the customized Lockbox LIMS was verified by Third Wave Analytics personnel to ensure that all customizations function appropriately per the unique user requirements for the SARS-CoV-2 detection workflow. To that end, all requirements were documented in Third Wave Analytics’ internal software verification system, first detailing all user requirements for how the system should function for the end laboratory user. Secondly, each user requirement then had a list of all the functional requirements for how the LIMS was specifically customized to meet the user requirements. Test cases were then created and run for each specific functional requirement, to document that each specific functional requirement was met. Upon the verification of the customized Lockbox LIMS software by Third Wave Analytics, the end users in the laboratory performed additional user acceptance testing to verify that the software performed as anticipated.

#### Automated Assay

Our fully automated assay is being established and validated at the time of this publication. We will update this manuscript to include our automated method when this workflow is implemented.

## Discussion and Conclusions

Here we provide a description of our path to establishing the IGI SARS-CoV-2 Diagnostic Testing Laboratory and the development and validation of a SARS-CoV-2 diagnostic test. Our RT-qPCR-based test shows a reproducible limit of detection of 1 × 10^3^ viral genomic copies/ml, comparing favorably to other validated tests in current use for community testing. We encountered several bottlenecks to converting an academic research laboratory into a clinical diagnostic testing facility, and we share our experience here to help others with this process. First and foremost, the laboratory must ensure that it meets the regulatory requirements for infrastructure and staff, including CLIA certification, HIPAA compliance, and FDA authorization. In our experience, extending the license from an existing CLIA certified and licensed facility, using online HIPAA training and adapting a commercially available FDA certified test for use with our equipment saved time and resources. In addition, we managed supply chain bottlenecks by using donated equipment, reducing the test-sample volume used in our assay, adopting in-house sample barcoding, and adapting materials such as sampling tubes to work with available equipment. To identify local sources of PPE, we posted a call for masks, gloves, and gowns on our website and succeeded in locating sufficient supplies for personnel use during the planned weeks of operation. Lab and supporting personnel were assigned to work in shifts and a succession plan is in place to ensure uninterrupted operation. In our experience, patient sample collection is the rate-limiting step in our sample analysis pipeline. We continue to address challenges of interfacing with health centers for patient sample intake and we will soon implement a web portal for this purpose that will be described in a future update to this manuscript.

We invested ∼$550,000 to cover initial costs of equipment and supplies. Our cost analysis indicates a per-test rate of $24 per test in reagents and consumables, and does not include personnel costs. We continue to investigate mechanisms of reimbursement including from local, state, and federal government sources. Our facility has also benefited from philanthropic donations to cover costs of tests for uninsured patients and for repetitive and asymptomatic patient testing.

As the COVID-19 pandemic continues, our intention is to provide both PCR-based diagnostic testing and to advance research on asymptomatic transmission, analyze virus sequence evolution, and provide benchmarking for new diagnostic technologies. If resources permit, we intend to conduct routine surveillance testing to enable rapid identification of infected individuals that will enable restriction of virus spread. In this way, academic testing centers like ours can limit viral transmission locally and enable the economy to restart nationally.

## Methods

FDA-approved diagnostic testing for COVID-19 infection is primarily based on quantitative real-time polymerase chain reactions (RT-qPCR) to measure SARS-CoV-2 viral RNA present within a patient specimen^6^. To do this, an intact specimen is collected by nasopharyngeal or oropharyngeal swab at a health care facility and is submitted to the diagnostic testing facility. RNA is extracted from the sample then RT-qPCR is used to detect viral S-gene, N-gene, and ORF1ab, along with an MS2 RNA technical control. Tests are scored as positive or negative for viral infection based on our validated limit of detection Ct threshold of 37, and using EUA approved criteria identified in Thermo Fisher’s instructions (Table 2).

### TEST VALIDATION

#### Limit of Detection

A serial dilution of the SARS-CoV-2 positive control RNA in the ThermoFisher TaqPath COVID-19 Control Kit was prepared in our sample collection medium (2X DNA/RNA Shield diluted to 1X in phosphate-buffered saline (PBS)), spanning 1 × 10^2^ to 5 × 10^4^ genomic copies of SARS-CoV-2 RNA per ml. The RT-qPCR reaction was run using triplicates at each concentration. The threshold values were set manually based on background fluorescence of the reaction mixture. MS2 bacteriophage genomic RNA (Thermo Fisher) was used as an internal control for RNA extraction, reverse transcription, and PCR amplification. We then performed 20 replicates at our target LoD to confirm the limit of detection.

#### Clinical Sample Evaluation Assay

##### Contrived samples and local testing facility samples

For the 30 positive samples, 24 mock positive samples we created by diluting (in a range near our LoD) Thermo Fisher’s SARS-CoV-2 positive control RNA from the TaqPath COVID-19 Control Kit into our sample collection medium (1X DNA/RNA Shield in PBS). 27 Mock negative samples were created by diluting RNA purified from cultured human 293T cells into IGI’s sample collection buffer (1X DNA/RNA Shield in PBS). Human RNA concentration was calculated using UV absorbance, and the concentration was converted to an approximate copy number for comparison with positive SARS-CoV-2 RNA controls using the NEBioCalculator for ssRNA with an estimated average RNA fragment length of 100nt.

Samples from the local testing facility arrived at the IGI in a frozen state, as 200 μl of leftover clinical samples already inactivated in DNA/RNA Shield. Since the local testing facility receives samples in UTM, they added equal volumes of 2X DNA/RNA Shield to inactivate their samples prior to entering their testing pipeline. We received 200 μl of these leftover inactivated samples. After thawing at IGI, sample collection medium (1X DNA/RNA Shield in PBS) was added to the 200 μl-samples to bring the final volume to 450 μl. These were then manually pipetted into 96-deep-well plates by transferring the full 450 μl, and RNA extraction and PCR reaction preparation were executed following the instructions in the *Semi-Manual SOP* (see supplementary files).

##### Kaiser Permanente Clinical Specimen Preparation

Ten clinical samples known to be positive for SARS-CoV-2 and ten clinical samples known to be negative for SARS-CoV-2 were obtained from Kaiser Permanente. These samples arrived in COPAN clinical specimen tubes, frozen in universal transport medium. These samples were received into the UC Berkeley BSL-3 facility by Dr. Sarah Stanley’s laboratory. In the BSL-3, samples were thawed and 450 μl of sample was mixed with 450 μl 2X DNA/RNA Shield to reach a final concentration of 1X DNA/RNA Shield. This sample (900 μl) was then transferred from the primary tube into a new sample tube in a BSL-2 biosafety cabinet within the BSL-3, and the primary tube was discarded into BSL-3 waste. The outside of the new sample tube was decontaminated and moved out of the BSL-3 for analysis at the IGI. Once at the IGI, the *Semi-Manual SOP* (see supplementary files) was followed to array the full contents (450 μl) of the clinical samples into 96 deep-well plates using the Hamilton STARlet, and procedures for manual pipetting of RNA extraction and RT-qPCR were followed exactly.

### PATIENT SAMPLE TESTING

While establishing a fully automated method, we have started testing patient samples with a semi-manual method at a lower scale. A detailed standard operating procedure for our semi-manual approach can be found in the supplementary file, *Semi-Manual SOP*. Additionally, a visual guide on the most important details of each step can be found in the Supplementary Figure 1. Notably, steps of laboratory testing (*e*.*g*., step start and completion time) are directly tracked by laboratory technicians using tablet computers that directly report to the LIMS.

#### Sample collection kit preparation

Sample collection kits are prepared in a separate laboratory from the testing facility. Kits are composed of an oropharyngeal swab, a short COPAN-like tube (ordered from IMPROVE medical) pre-filled with the IGI’s sample collection medium (sterile 1X DNA/RNA Shield in PBS), in a zip-top biohazard bag. Lot numbers for preparing and pre-loading tubes with sample collection medium are recorded on the bags. At the Tang Center, patient intake produced barcodes that are affixed to the sample collection tube and intake paperwork and the sample is returned to IGI in a secondary container at ambient temperature. Details on kit assembly, and patient sample collection and transport at the Tang Center can be found in the supplementary file, *Kit Preparation and Patient Sample Collection and Transport*.

#### Acquisition

Briefly, during sample acquisition, a kit is received with a patient sample tube and biohazard bag containing matching barcodes and patient identifying information. The kits are then examined to determine acceptance or rejection for testing. Rejection criteria include: fully or partially uncapped sample tube, absence of swab, compromised sample identification (illegible or tube identification not matching the biohazard bag), observable contamination of collection medium, and visible liquid in the kit’s biohazard bag. For rejected sample kits, the disposal will be made in biohazard waste and recollection from the respective patients under investigation will be pursued by our health partners when possible.

An accepted patient sample tube is sprayed with 70% molecular biology-grade ethanol, wiped down with a Kimwipe in the biosafety cabinet and stored at 4°C or plugged into the testing workflow. The testing workflow consists of four main steps: Accessioning, RNA Extraction, RT-qPCR and Resulting.

#### Accessioning

The clean barcoded patient sample tube is placed in a Hamilton Microlab STARlet rack to be scanned and arrayed into a barcoded 96-deep-well plate, which will then either be stored at 4°C or transferred directly to an extraction personnel for the RNA Extraction phase. Please see the supplementary file, *Hamilton Microlab STARlet Automation Process*, for details on the STARlet process and our bespoke STARlet automation code. The STARlet output .csv file is uploaded into the LIMS to register the now de-identified samples into the database as received and pending extraction.

#### RNA Extraction

Extraction personnel place the centrifuged and sealed deep-well plates into the PCR clean hood for RNA extraction. In the hood, the reagents are prepared for the Thermo Fisher EUA MagMAX Viral/Pathogen Nucleic Acid Isolation Kit using our reduced reaction volume approach (see *Validation Methods* section), and samples are processed by digestion with proteinase K and addition of nucleic acid-binding magnetic beads. Pipette tips should be switched between samples throughout the process to avoid cross-contamination. MS2 genomic RNA (from Thermo Fisher’s TaqPath RT-PCR COVID-19 Kit) is spiked into every well as an internal control for RNA extraction and RT-PCR. Negative controls are a 1:1 mix of 2X DNA/RNA Shield in phosphate-buffered saline (PBS) and human cell line RNA. Using the LIMS interface, extraction personnel manually indicate completion of each step in the RNA extraction protocol. The RNA extraction plate is either transferred directly to RT-qPCR personnel or stored at 4°C for 1h, −20°C overnight or −80°C indefinitely.

#### RT-qPCR

For RT-qPCR, the TaqPath RT-PCR COVID-19 Kit (Thermo Fisher) COVID-19 positive control RNA at 1 × 10^4^ genomic copies/ml is used. Negative control is molecular biology grade water (Thermo Fisher or equivalent). RT-qPCR personnel thaw pre-loaded master mix plates from −20°C, out of the light, for 10 min. Replicate RT-qPCR plates are prepared with matching sample positions. Identification of RT-qPCR plates used are manually entered in the LIMS interface and RT-qPCR steps are manually checked as they are completed. The PCR is held in an Applied Biosystems 7500 Fast ThermalCycler (Thermo Fisher). Reaction outputs for both plates are manually uploaded to the LIMS system for analysis.

#### Resulting

The uploaded outputs are analyzed and synthesized in the LIMS. Negative and positive results are evaluated by a CLS in the LIMS. A new specimen will be requested for any samples that do not result in a negative or positive test result after two full tests. Possible reported results include: Positive”, “negative”, “new specimen requested”. If sample results are discordant between samples on replicate plates, or if result is invalid or inconclusive the sample is requeued by the LIMS for a full retest starting in the accessioning phase (Table 2).

Upon a valid result and review by the CLS, results need to be reported to the relevant physicians as well as state and local authorities. Our resulting workflow is as follows: 1) call the physician that requisitioned the test with any critical (positive) results, 2) return all “positive,” “negative,” “inconclusive,” or “specimen insufficient” results as a PDF report per patient to the physician, then 3) after the test results have been officially reported, all “positive,” “negative,” and “inconclusive” results are transmitted as a .csv in a HIPAA-compliant manner (using UC Berkeley’s Google email service with additional Virtru encryption) to CDPH to be uploaded to the California Reportable Disease Information Exchange (CalREDIE). Typically, reporting to local health authorities must be done within one hour of reporting a positive result to a clinician in a format specified by the public health authority and reporting to CDPH must be done within 24 hours via electronic transmission.

However, public health orders from the Bay Area jurisdictions served by our clinical lab called for reporting of all COVID-19 test results (not just positives). These same orders by local public health authorities further specified that electronic transmission of results to the CalREDIE system fulfilled reporting requirements to them, obviating the need to report directly to Bay Area local health authorities for COVID-19 testing. CDPH uses CalREDIE to then report on state-level statistics to the CDC. In phase two of the LIMS implementation, the physician access portal will provide results in PDF format to the requisitioning physician, integrate with provider electronic medical record systems like EPIC, and return CDPH reports to CalREDIE via an HL7 interface instead of .csv.

#### LIMS Development

The Lockbox LIMS software v1.48 was used as the template for the LIMS, which is an OEM installed software package run on the Salesforce platform. Significant customizations were implemented to meet all the laboratory user requirements of the SARS-CoV-2 detection workflow. These customizations included: 1) automated sample record creation upon upload of sample barcodes, 2) the display of customized sample processing instructions for the laboratory user as samples are tested (e.g., sample testing SOPs), 3) automated record updating throughout the sample testing process (e.g., automated updating of sample or testing plates statuses), 4) creation of user-friendly data import tools (e.g., upload of raw Ct data upon completion of RT-qPCR), 5) automated analysis of all control and sample data to generate a final patient result instantaneously upon raw data upload, 6) customized and user-friendly workflow for the final review of sample data by appropriate laboratory personnel, 7) export of the result data for import into health partners systems (e.g. the UHS Tang Center), 8) automated sample record creation upon the re-testing of a sample, 9) the tracking of all reagents and control lots during testing, 10) customized dashboards to track all samples throughout the testing process, 11) creation of a provider portal (in the form of a partner community) for the ordering and reporting of results to non-UC Berkeley patients, and 12) the implementation of robust security measures to ensure that all PHI are encrypted (at rest) and that only appropriate personnel have access to appropriate records and data fields.

### INSTITUTIONAL APPROVAL FOR HUMAN SUBJECT DATA

Descriptive statistics for patient samples used in this manuscript come from de-identified datasets in accordance with human subjects protections, as approved by UC Berkeley’s Committee for Protection of Human Subjects (CPHS). While Institutional Review Board (IRB) approval is required for any research using human subjects, clinical laboratory activities exclusively supporting CLIA-certified clinical operations do not. These activities are governed by CMS and HIPAA legislation. In order to publish our data in this manuscript as results of developing a testing workstream, we sought IRB approval through UC Berkeley’s CPHS. The UC Berkeley Committee for Protection of Human Subjects determined that all the analyses presented in this manuscript do not qualify as human subjects research as the data sets were de-identified to those analyzing them for these results (IRB submission # 2020-04-13177).

## Data Availability

All data are available in figures, supplemental files and linked materials.

https://innovativegenomics.org/sars-cov-2-testing-guide/

## Acknowledgments

We are indebted to the following organizations and individuals whose support and contributions made our work possible:

UC Berkeley Chancellor Carol Christ, UC Berkeley Vice Chancellor for Research Randy Katz, UC Berkeley Dean for Biomedical Sciences Michael Botchan, lead UC Counsel Charles Robinson, and UC President Janet Napolitano provided strategic support throughout the course of our effort.

Prof. Robert Tjian, MCB Department, enabled our effort by consistent engagement and action at every level necessary.

UC Berkeley Vice-Chancellor for Health, Dr. Guy Nicolette, Holly Stern and the Tang Center chief medical officer, Dr Anna Harte, enabled and guided our interface to University Health Services.

Prof. Joseph DeRisi and Dr. Eric Chow (CZ Biohub) allowed our personnel to visit the CZ Biohub where UCSF Clinical operates a SARS-CoV-2 diagnostic laboratory and shared key details about their organizational setup and workflow that informed our own effort.

Dr. Lisa Hernandez (City of Berkeley Public Health) and Chief David Brannigan (City of Berkeley Fire Department) envisioned and led the establishment of a partnership between the City, the IGI, and LifeLong Medical. Inspector Dori Tieu (City of Berkeley Fire Department) provided key logistical and operational support.

Dr. Michael Stacey, Dr. Kim Nguyen, and Yui Nishiike (LifeLong Medical Care) provided key guidance and support on establishing workflows at the IGI-LifeLong interface.

Dr. Jeffrey Shapiro and Dr. Jacek Skarbinsky (Kaiser Permanente) provided de-identified patient samples.

Drs. Alan Sachs, Elizabeth Kerr, Christopher Cowen, and David Woo (Thermo Fisher) provided scientific and logistical support and ensured a sustained supply chain.

Drs. Ray Turner and Nigel Mouncey (JGI) provided enabling personnel support.

The Dillin, Karpen, Naar, Niyogi, and Herr laboratories donated essential lab equipment.

Pattern and UC Berkeley College of Engineering donated PPE.

Thomson Reuters provided our staff with HIPAA training.

Initial funding was provided by IGI Founders Fund, Dr. Anders Naar, Rick and Rachel Klausner, and the Shurl and Kay Curci Foundation.

We also thank Dr. Jamie Cate, UC Berkeley CEND Director Dr. Julia Schaletsky, Hamilton, Bayer, Salesforce, ThirdWave Analytics, Illumina, Active Motif, Riffyn, ACT Catering, USA Scientific, Sartorius, NanoCellect, Dovetail Genomics, Takara Bio, Immudex, Twist Bioscience, Fisher Scientific, 10X Genomics, and Novogene for support, encouragement, and sustenance!

## Contact

You can reach out to us at with any questions or comments.

### Commonly-Used Abbreviations

FDA: Food and Drug Administration
CDC: Centers for Disease Control and Prevention
CMS: Centers for Medicare & Medicaid Services
CDPH: California Department of Public Health
LFS: Laboratory Field Services (division of CDPH that regulates CLIA)
LDT: Laboratory-Developed Test
CLIA: Clinical Laboratory Improvement Amendments
EUA: Emergency Use Authorization (under FDA)
RT-qPCR: Quantitative Reverse Transcription Polymerase Chain Reaction
LIMS: Laboratory Information Management System

## Supplementary Material

### USEFUL LINKS

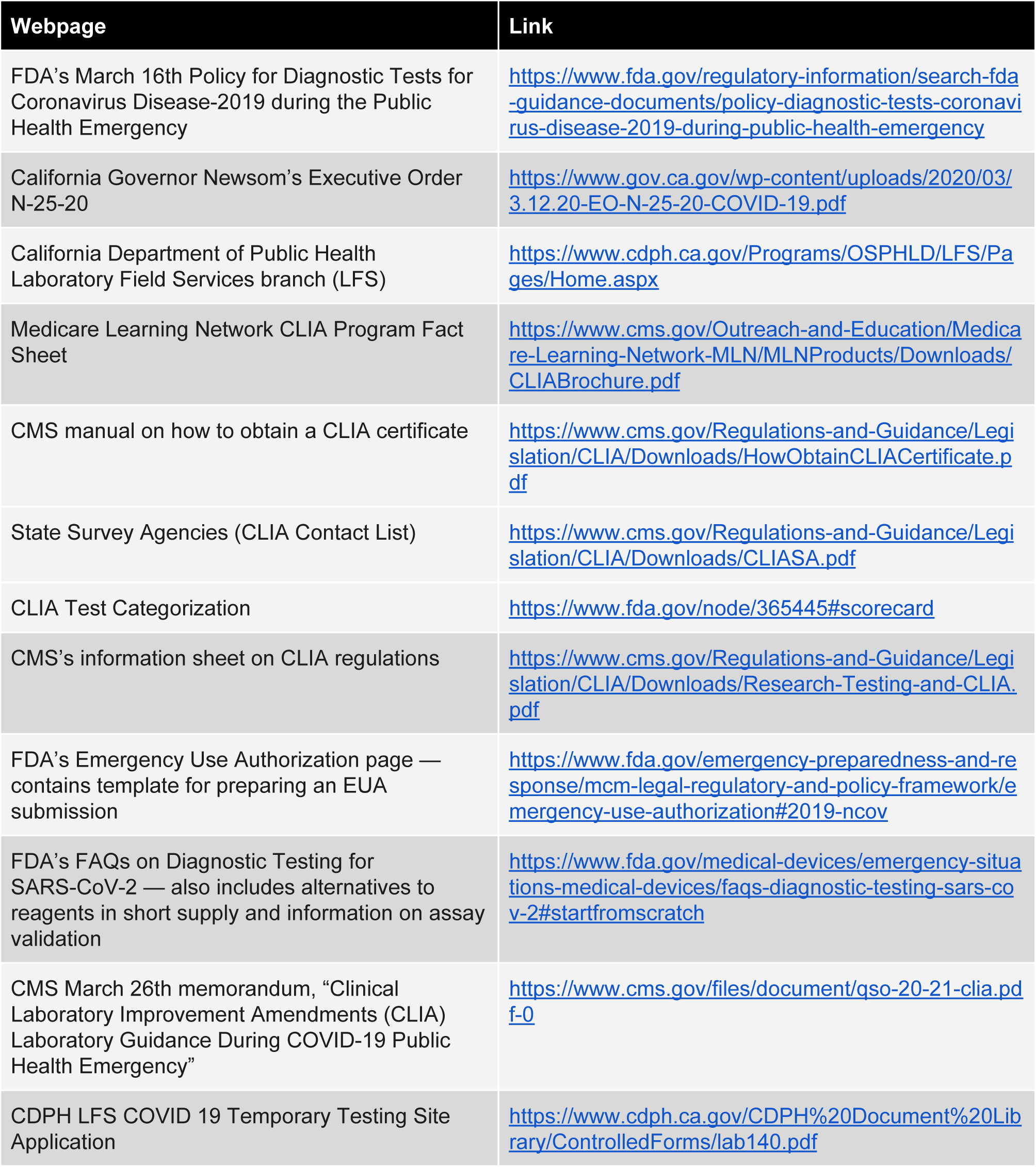

| Webpage | Link |
| --- | --- |
| FDA's March 16th Policy for Diagnostic Tests for Coronavirus Disease-2019 during the Public Health Emergency | https://www.fda.gov/regulatory-information/search-fda-guidance-documents/policy-diagnostic-tests-coronavirus-disease-2019-during-public-health-emergency |
| California Governor Newsom's Executive Order N-25-20 | https://www.gov.ca.gov/wp-content/uploads/2020/03/3.12.20-EO-N-25-20-COVID-19.pdf |
| California Department of Public Health Laboratory Field Services branch (LFS) | https://www.cdph.ca.gov/Programs/OSPHLD/LFS/Pages/Home.aspx |
| Medicare Learning Network CLIA Program Fact Sheet | https://www.cms.gov/Outreach-and-Education/Medicare-Learning-Network-MLN/MLNProducts/Downloads/CLIABrochure.pdf |
| CMS manual on how to obtain a CLIA certificate | https://www.cms.gov/Regulations-and-Guidance/Legislation/CLIA/Downloads/HowObtainCLIACertificate.pdf |
| State Survey Agencies (CLIA Contact List) | https://www.cms.gov/Regulations-and-Guidance/Legislation/CLIA/Downloads/CLIASA.pdf |
| CLIA Test Categorization | https://www.fda.gov/node/365445#scorecard |
| CMS's information sheet on CLIA regulations | https://www.cms.gov/Regulations-and-Guidance/Legislation/CLIA/Downloads/Research-Testing-and-CLIA.pdf |
| FDA's Emergency Use Authorization page — contains template for preparing an EUA submission | https://www.fda.gov/emergency-preparedness-and-response/mcm-legal-regulatory-and-policy-framework/emergency-use-authorization#2019-ncov |
| FDA's FAQs on Diagnostic Testing for SARS-CoV-2 — also includes alternatives to reagents in short supply and information on assay validation | https://www.fda.gov/medical-devices/emergency-situations-medical-devices/faqs-diagnostic-testing-sars-cov-2#startfromscratch |
| CMS March 26th memorandum, "Clinical Laboratory Improvement Amendments (CLIA) Laboratory Guidance During COVID-19 Public Health Emergency" | https://www.cms.gov/files/document/qso-20-21-clia.pdf |
| CDPH LFS COVID 19 Temporary Testing Site Application | https://www.cdph.ca.gov/CDPH%20Document%20Library/ControlledForms/lab140.pdf |

## SUPPLEMENTARY FILES

*For links to all supplementary files and high-resolution supplementary figures, please visit:* https://innovativegenomics.org/sars-cov-2-testing-guide/

**Supplementary Figure 1 (part 1 of 2).**
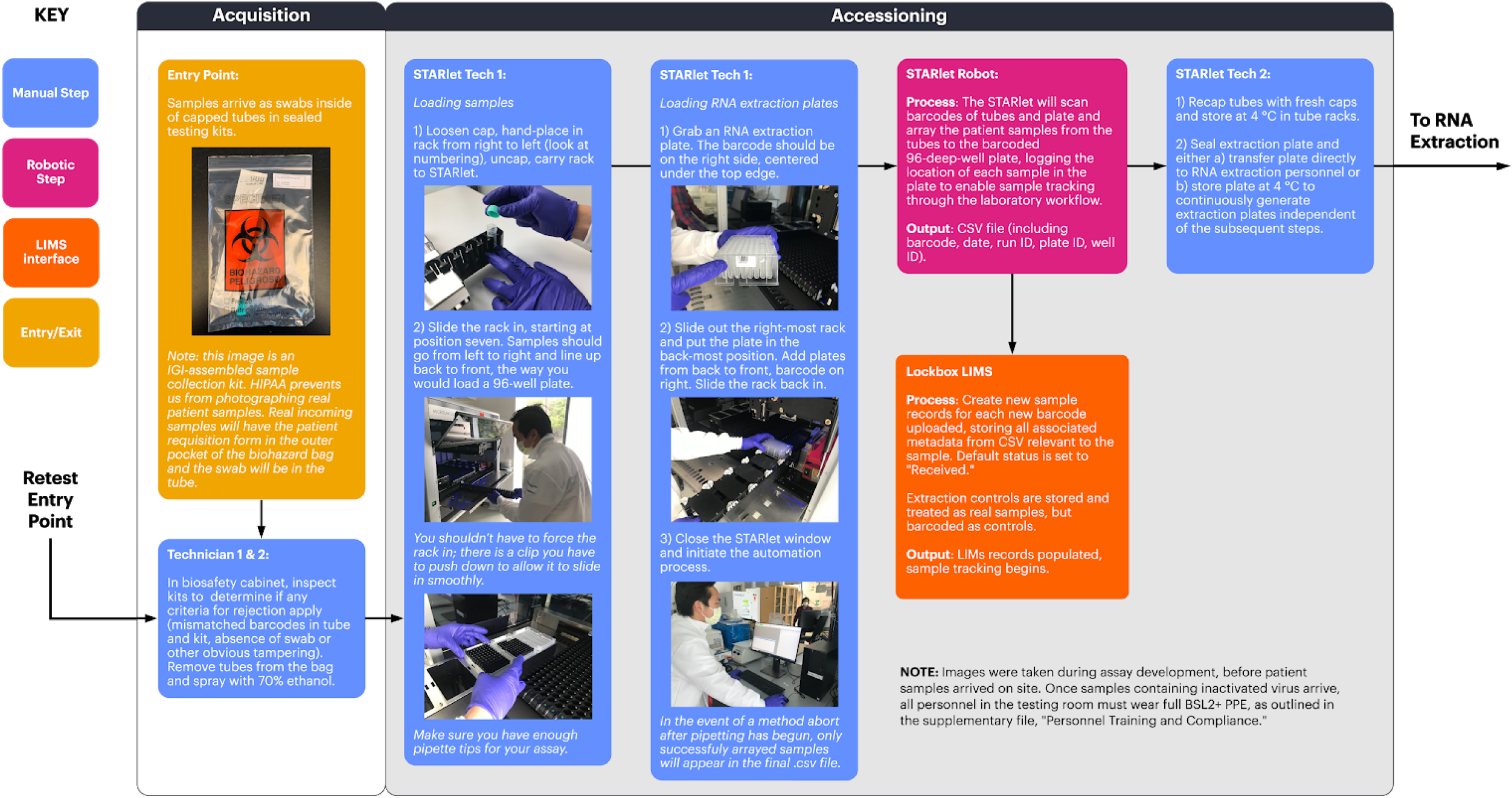
Illustrated diagram of our semi-manual workflow, from acquisition to accessioning. Click [here](https://berkeley.box.com/s/e3udaly3mj8zhg3zvugd7amovugdfo9b) for combined, high-res figure.

**Supplementary Figure 1 (part 2 of 2).**
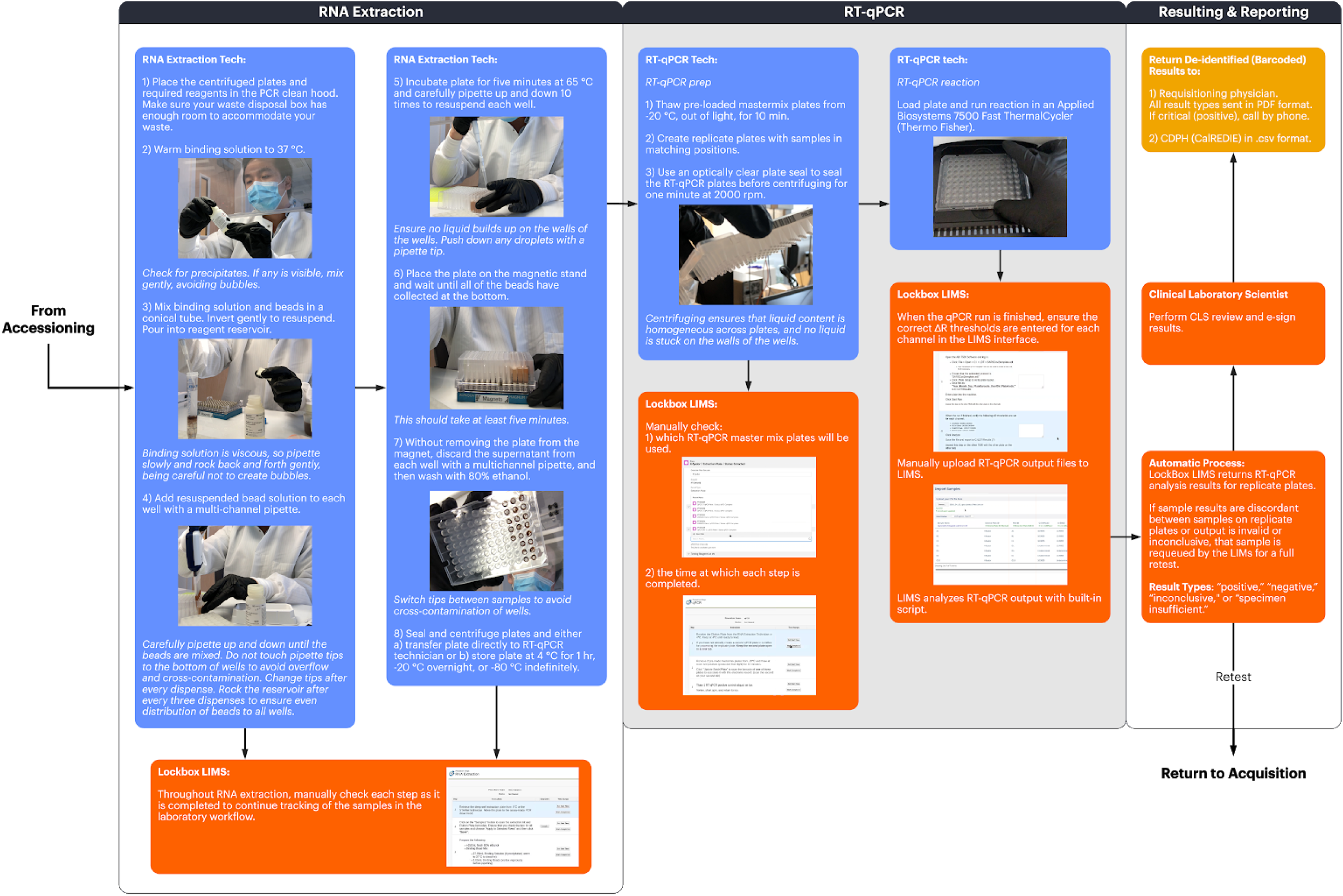
Illustrated diagram of our semi-manual workflow, from RNA extraction to resulting and reporting. Click [here](https://berkeley.box.com/s/e3udaly3mj8zhg3zvugd7amovugdfo9b) for combined, high-res figure.

**Supplementary Figure 2.**
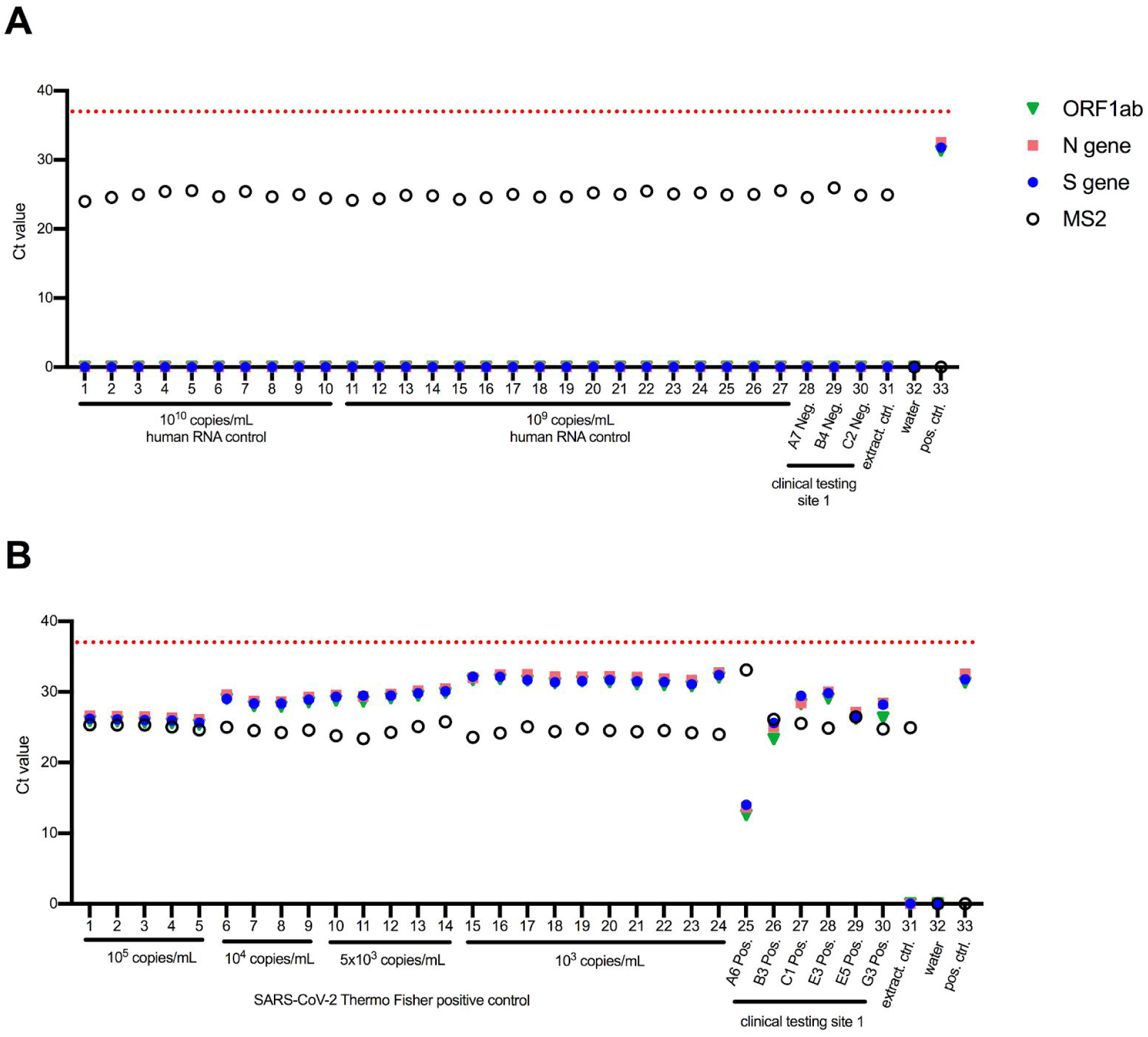
Clinical Validation, Contrived and Clinical Samples - one of duplicate PCR plates. The results from the corresponding duplicate PCR plate are shown in Figure 4. Replicates are plotted as individual points (rather than mean values). An undetermined Ct value is plotted as Ct = 0. **A**: Contrived negative and local testing facility known negative samples (clinical testing site 1). Whole cell RNA purified from cultured human 293T cells was diluted in IGI’s sample collection buffer (1X DNA/RNA Shield in PBS) to approximate negative samples. Human RNA concentration was calculated using UV absorbance, and the concentration was converted to an approximate copy number for comparison with positive RNA controls (See methods for details). Assay quality controls are shown in lanes 31-33, representing the extraction control (Thermo Fisher’s MS2 control in 1X DNA/RNA shield and PBS), and two PCR controls - water, and the Thermo Fisher kit positive SARS-CoV-2 RNA control (without MS2 RNA). **B**: Contrived positive and local testing facility known positive samples. Contrived positive samples were prepared by diluting SARS-CoV-2 positive control RNA from Thermo Fisher’s kit into IGI’s sample collection buffer (1X DNA/RNA Shield in PBS). Assay quality controls are the same as in B.

**Supplementary Figure 3.**
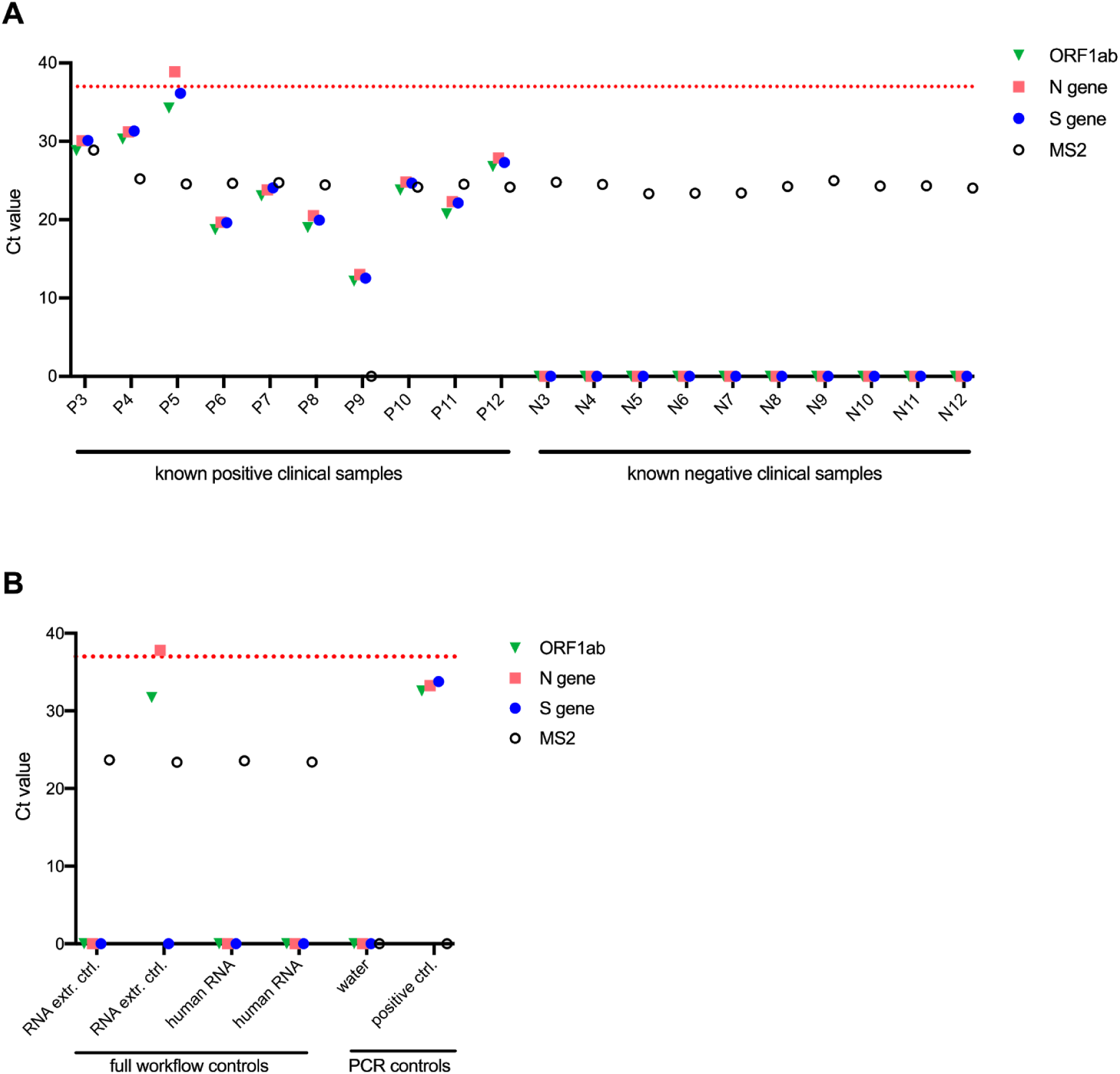
Clinical validation assay using leftover samples from Kaiser Permanente - one of duplicate PCR plates. The results from the corresponding duplicate PCR plate are shown in Figure 5. Undetermined Ct values are plotted as Ct = 0. **A**: Ten known positive and ten known negative patient samples in UTM leftover from Kaiser Permanente’s clinical testing facility were delivered to Sarah Stanley’s BSL-3 lab at UC Berkeley where they were inactivated by diluting 1:1 in 2X DNA/RNA Shield. Decontaminated tubes were delivered to IGIB where they entered our testing pipeline, starting with arraying into 96-deep-well plates using the Hamilton STARlet robot. **B**: Assay quality controls. Full workflow controls were processed starting at RNA extraction in the 96-well plates. RNA extraction control is composed of Thermo Fisher’s MS2 control in 1X DNA/RNA shield and PBS. Human RNA controls are composed of 200 ng/ml purified RNA from 293T cells in 1X DNA/RNA shield and PBS. PCR controls were processed starting at assembling RT-qPCR reactions and include a water control, and the Thermo Fisher kit positive SARS-CoV-2 RNA control (without MS2). The second RNA extract control from the left returned two high Ct values and one undetermined value for SARS-CoV-2. Based on our decision matrix criteria for resulting (Table 2), this control returns a NEG result (positive for only 1 SARS-CoV-2 gene). While this meets our acceptance criteria, we expect that there may have been some cross contamination from neighboring wells during sealing and unsealing of the plate during an unexpected instrument calibration step.

## References

1. World Health Organization. Coronavirus disease (COVID-19) Pandemic. (2020).

2. Johns Hopkins University. Coronavirus COVID-19 Global Cases by the Center for Systems Science and Engineering (CSSE). (2020).

3. Center for Disease Control. Implementation of Mitigation Strategies for Communities with Local COVID-19 Transmission. (2020).

4. Li, R. et al.. Substantial undocumented infection facilitates the rapid dissemination of novel coronavirus (SARS-CoV2). Science (2020). doi:10.1126/science.abb3221

5. Wu, Z. & McGoogan, J. M. Characteristics of and Important Lessons From the Coronavirus Disease 2019 (COVID-19) Outbreak in China. JAMA (2020). doi:10.1001/jama.2020.2648

6. Guan, W.-J. et al. Clinical Characteristics of Coronavirus Disease 2019 in China. N. Engl. J. Med.(2020). doi:10.1056/NEJMoa2002032

7. The Economist, Science and Technology, April 2 Edition: https://www.economist.com/science-and-technology/2020/04/02/an-antibody-test-for-the-novel-coronavirus-will-soon-be-available

8. Bennhold, K. “A German Exception? Why the Country’s Coronavirus Death Rate Is Low.” The New York Times April 4, 2020 Web. 9 March 2020 https://www.nytimes.com/2020/04/04/world/europe/germany-coronavirus-death-rate.html

9. Şimşek, A. “COVID-19: Germany deaths hit 815, infection rate slows; Lockdown measures, widespread testing slowing spread of coronavirus in Germany.” Anadolu Agency. 1 April 2020. Web. 9 March 2020 https://www.aa.com.tr/en/europe/covid-19-germany-deaths-hit-815-infection-rate-slows/1788307.

10. Food and Drug Administration. FAQs on Diagnostic Testing for SARS-CoV-2. (2020).

11. Applied Biosystems. TaqPath COVID-19 Combo Kit Instructions for Use. Publication Number MAN0019181, Revision B.0. (2020)

12. Thermo Fisher Scientific. TaqPath COVID-19 Multiplex Diagnostic Solution. Web. 10 March 2020. https://www.thermofisher.com/us/en/home/clinical/clinical-genomics/pathogen-detection-solutions/coronavirus-2019-ncov/genetic-analysis/taqpath-rt-pcr-covid-19-kit.html

